# Rapid implementation of real-time SARS-CoV-2 sequencing to investigate healthcare-associated COVID-19 infections

**DOI:** 10.1101/2020.05.08.20095687

**Authors:** Luke W. Meredith, William L. Hamilton, Ben Warne, Charlotte J. Houldcroft, Myra Hosmillo, Aminu S. Jahun, Martin D. Curran, Surendra Parmar, Laura G. Caller, Sarah L. Caddy, Fahad A. Khokhar, Anna Yakovleva, Grant Hall, Theresa Feltwell, Sally Forrest, Sushmita Sridhar, Michael P. Weekes, Stephen Baker, Nicholas Brown, Elinor Moore, Ashley Popay, Iain Roddick, Mark Reacher, Theodore Gouliouris, Sharon J. Peacock, Gordon Dougan, M. Estée Török, Ian Goodfellow

**Author notes:** These authors contributed equally to the work (Luke W. Meredith, William L. Hamilton, M. Estée Török, Ian Goodfellow). Joint first author (Luke W. Meredith, William L. Hamilton). Joint last author (M. Estée Török, Ian Goodfellow). **Corresponding Authors:** Dr M. Estée Török, Professor Ian Goodfellow.

## Abstract

**Background:** The burden and impact of healthcare-associated COVID-19 infections is unknown. We aimed to examine the utility of rapid sequencing of SARS-CoV-2 combined with detailed epidemiological analysis to investigate healthcare-associated COVID-19 infections and to inform infection control measures.

**Methods:** We set up rapid viral sequencing of SARS-CoV-2 from PCR-positive diagnostic samples using nanopore sequencing, enabling sample-to-sequence in less than 24 hours. We established a rapid review and reporting system with integration of genomic and epidemiological data to investigate suspected cases of healthcare-associated COVID-19.

**Results:** Between 13 March and 24 April 2020 we collected clinical data and samples from 5191 COVID-19 patients in the East of England. We sequenced 1000 samples, producing 747 complete viral genomes. We conducted combined epidemiological and genomic analysis of 299 patients at our hospital and identified 26 genomic clusters involving 114 patients. 66 cases (57.9%) had a strong epidemiological link and 15 cases (13.2%) had a plausible epidemiological link. These results were fed back to clinical, infection control and hospital management teams, resulting in infection control interventions and informing patient safety reporting.

**Conclusions:** We established real-time genomic surveillance of SARS-CoV-2 in a UK hospital and demonstrated the benefit of combined genomic and epidemiological analysis for the investigation of healthcare-associated COVID-19 infections. This approach enabled us to detect cryptic transmission events and identify opportunities to target infection control interventions to reduce further healthcare-associated infections.

## Introduction

SARS-CoV-2 emerged in the human population in December 2019^1^, originating from an intermediate animal host^2^. Owing to the error prone nature of the viral replication process, RNA viruses, such as SARS-CoV-2, accumulate mutations over time which results in sequence diversity. The current mutation rate of SARS-CoV-2 is estimated to be ~2.5 nucleotides per month^3,4^. Sequencing of SARS-CoV-2 can provide valuable information on virus biology, transmission and population dynamics^5–8^. When linked with detailed epidemiological data and performed in a timescale of days, genomic data can support epidemiological investigations of potential hospital-acquired infections (HAI). On a larger population scale, genomic surveillance of SARS-CoV-2 can inform which lineages of the virus are currently circulating in the human population, how these change over time as an indicator of the success of control measures, how often new sources of virus are introduced from other geographic areas, and how the virus evolves in response to interventions.

Healthcare-associated infections (HCAI) can affect patients, resulting in increased morbidity and mortality, and healthcare workers (HCW), impacting on staff sickness and morale, to the detriment of patient care. It is crucial to rapidly detect and manage HCAI effectively to prevent both complications and onward transmission to susceptible patients and staff. The burden of nosocomial COVID-19 infections is unknown but one study from China has reported a 41% prevalence^9^ Worldwide, over 22,000 cases of COVID-19 infection in healthcare workers have been reported, but is likely to be an underestimate^10^. As the number of community-acquired COVID-19 cases reduces, healthcare settings are likely to act as reservoirs of infection. Identifying transmission events in these settings will therefore become increasingly important to manage outbreaks and monitor effective infection control.

We aimed to examine the utility of rapid sequencing of SARS-CoV-2, combined with detailed epidemiological analysis, to investigate healthcare-associated COVID-19 infections and to inform infection control measures in our hospital.

## Methods

### Study design, setting and participants

A prospective surveillance study of COVID-19 infections was conducted at Cambridge University Hospitals NHS Foundation Trust (CUH), a secondary care provider and tertiary referral centre in the East of England (EoE). Clinical specimens collected from patients presenting to 18 hospitals in EoE were submitted to the Public Health England Clinical Microbiology and Public Health Laboratory (CMPHL) at CUH for diagnostic testing. Samples underwent nucleic acid extraction and were tested for presence of SARS-CoV-2 using a validated in-house RT qPCR assay developed by CMPHL (Supplementary Methods). The test was reported as SARS-CoV-2 PCR positive if the cycle threshold (Ct) value was ≤36.

### Study procedures

Demographic, clinical and laboratory data were extracted from the hospital information system (EPIC Systems Corporation, Verona, USA). All PCR-positive diagnostic samples were identified and transferred from the CMPHL to the Department of Virology for nanopore sequencing. All CUH samples and a selection of samples from East of England hospitals were selected for sequencing.

### Sequencing

Where Ct values were available prior to sample selection, positive samples with a Ct value ≤33 were sequenced using a multiplex PCR based approach according to the modified ARTIC v2 protocol v3 primer set^11,12^. Amplicon libraries were sequenced using MinION flow cells v9.4.1 (Oxford Nanopore Technologies, Oxford, UK). Genomes were assembled using reference-based assembly and a bioinformatic pipeline^13^ using 20x minimum coverage cut-off for any region of the genome and 50.1% cut-off for calling single nucleotide polymorphisms.

### Genomic analysis

All sequences underwent quality control (QC) filtering and de-duplication to remove repeat samples from the same patient. Multiple sequence alignment was performed using MAFFT^14^. Phylogenetic trees were produced using IQ-TREE^15^ and visualised in Microreact^16^ for weekly hospital reports and the R package ggtree^17^. A pairwise SNP distance matrix was produced from the alignment (Supplementary Methods). Viral lineages^18^ were assigned with the PANGOLIN package (https://github.com/hCoV-2019/pangolin) v1.07.

### Epidemiological analysis

Patient movement data for all SARS-CoV-2 PCR positive samples were extracted from the hospital information system and transferred to the PHE Field Service (Epidemiology). Epidemiological analysis was performed using a cloud-based plotter application (Cluster Track, Camart Ltd, Cambridge, UK).

### Review of healthcare-associated infections

Clusters of COVID-19 cases including HCWs were identified by the clinical and infection control teams and reviewed at a weekly meeting, co-ordinated by the Patient Safety team. The genomic and epidemiological analyses were presented, to help determine whether infections were healthcare-associated and to identify possible causes and interventions. A weekly report was fed back to the clinical, infection control and hospital management teams to inform changes in infection control practice and comply with patient safety procedures.

### Ethical considerations

The study was conducted as part of surveillance for COVID-19 infections under the auspices of Section 251 of the NHS Act 2006. It therefore did not require individual patient consent or ethical approval. The COG-UK study protocol was approved by the Public Health England Research Ethics Governance Group.

## Results

### Baseline characteristics and COVID-19 epidemic curve

368 PCR-confirmed COVID-19 patients were admitted to CUH between 10 March and 24 April 2020 (Figure 1, Table 1). The median age was 66 years (range 0 to 98 years) and 63% were male. 19.6% of patients were admitted to critical care units and 20.1% died. Between 13 March to 24 April 2020 1000 samples were selected for sequencing and, after QC and de-duplication, 747 were used for downstream analysis (Supplementary Figure 1). 256/368 (69.6%) of CUH COVID-19 admissions had sequencing data available (Supplementary Table 1). 52/368 (14.1%) of admissions were suspected or highly likely hospital-acquired, of which 47 (90.4%) had genome sequences available. A further 10.3% of admissions were community-acquired but likely healthcare-associated, and 2.2% were healthcare workers (Supplementary Table 1). The CUH epidemic curve showed that weekly admissions peaked in week 4 (commencing 30 March 2020) and then declined (Figure 2). The UK went into full lockdown on 23 March 2020. In the early stages of the epidemic community-onset community-acquired infections predominated but the frequency of healthcare-associated infections increased over time.

**Figure 1.**
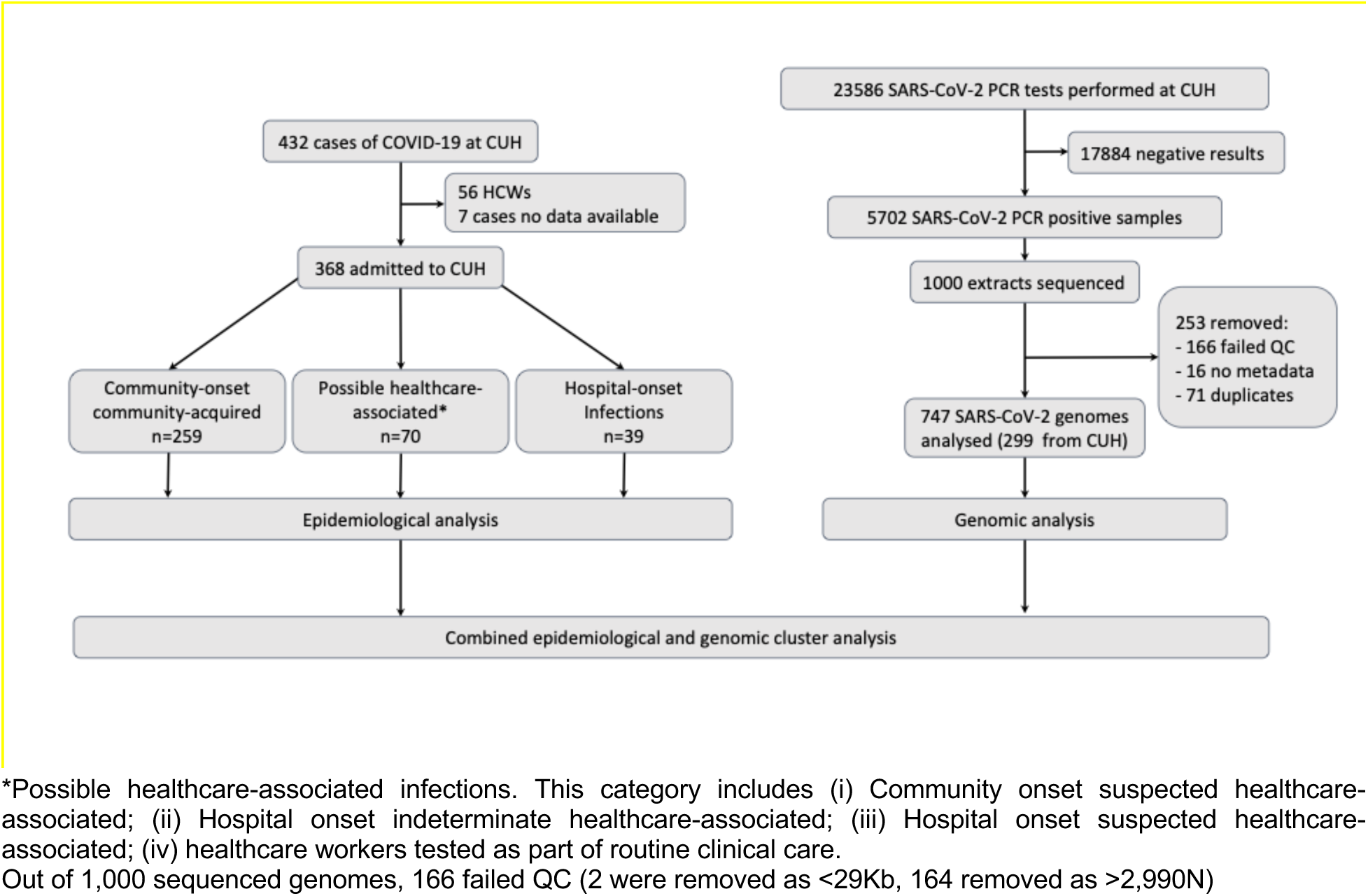
Study flow diagram.

**Figure 2.**
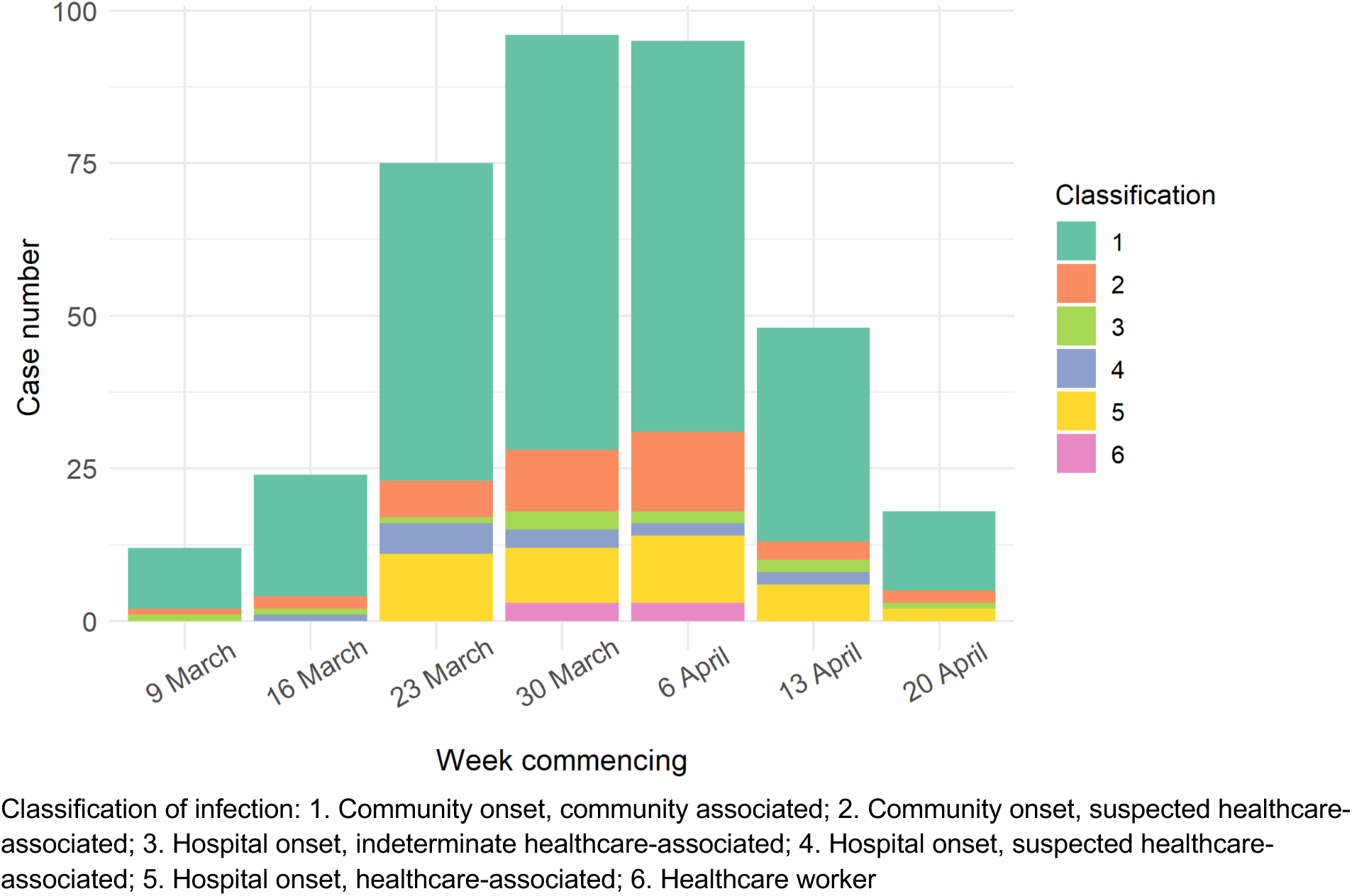
Epidemic curve of COVID-19 infections at CUH.

**Table 1.** Baseline characteristics of COVID-19 patients at CUH.

|  |  |
| --- | --- |
| **Baseline characteristics** |  |
| Number of patients | 374 |
| Age in years, mean (range)<br>Age in years, median (IQR) | 64 (0-98)<br>67 (51-79) |
| Male sex | 233 (62.3%) |
| Female sex | 141 (37.7%) |
| Ethnicity – White | 267 (71.4%) |
| Ethnicity – Black, Asian and minority ethnic | 28 (7.5%) |
| Ethnicity – not stated/missing | 79 (21.1%) |
| **Co-morbidities** |  |
| Hypertension | 115 (31.3%) |
| Ischaemic heart disease | 53 (14.4%) |
| Cardiac failure | 25 (6.8%) |
| Asthma | 45 (12.8%) |
| Chronic obstructive pulmonary disease | 30 (8.2%) |
| Diabetes mellitus | 78 (21.2%) |
| Chronic kidney disease | 37 (9.9%) |
| Hepatic cirrhosis | 16 (4.4%) |
| Dementia | 32 (8.7%) |
| Obesity | 51 (13.9%) |
| **Classification of infection\*** |  |
| Community onset, community associated | 261 (69.9%) |
| Community onset, suspected healthcare-associated | 40 (10.7%) |
| Hospital onset, indeterminate healthcare-associated | 11 (2.9%) |
| Hospital onset, suspected healthcare-associated | 13 (3.5%) |
| Hospital onset, healthcare-associated | 39 (10.5%) |
| Healthcare worker | 9 (2.4%) |
| Unknown | 1 (0.3%) |
| **Outcome** |  |
| Admission to hospital | 347 (92.5%) |
| Admission to critical care | 72 (19.3%) |
| Died as an inpatient | 75 (20.1%) |

### Genomic analysis

Each week a sample set was locked and underwent bioinformatic analysis. Out of 1,000 sequenced genomes, 253 were excluded from downstream analysis because they failed to pass QC thresholds, metadata were missing or they were repeat samples from the same patient (Figure 1). The median genome depth of coverage was 6,612x. We compared Ct value versus depth of coverage and found that the latter declined at Ct values >30 (Supplementary Figure 2). We also examined the location and frequency of SNPs across the genomes (Supplementary Figure 2). Genomes were assigned to a lineage based on the combination of mutations that have accumulated since the virus emerged. As of 23 March 2020, 12 lineages had been described in the UK^19^. The vast majority of samples in both the EoE and CUH belonged to lineage B.1. There were no lineage A samples, which have mainly been identified in China, USA, South Korea and Australia^18^ (Figure 3, Supplementary Figure 3).

**Figure 3.**
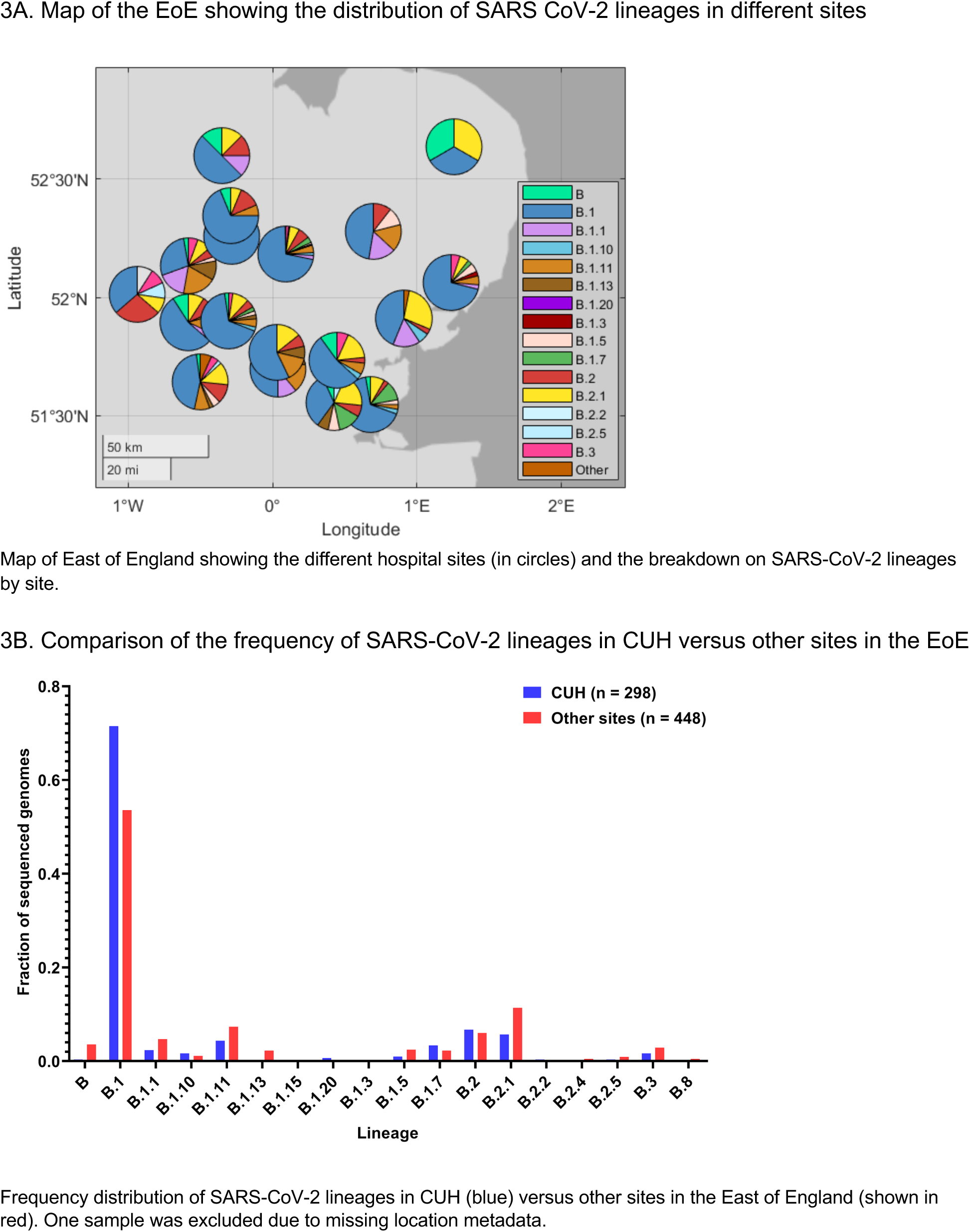
SARS-CoV-2 lineages identified in the East of England and CUH.

Phylogenetic trees were used to explore potential genetic clustering and correlation with sampling ward location and/or cases of suspected HAI (Figure 4). Samples collected from the Emergency Department (ED) were phylogenetically dispersed, likely reflecting unconnected recent transmission events in community-acquired infections (Supplementary Figure 4). In contrast, samples collected from several wards in CUH and an outpatient dialysis unit were genetically clustered (Figure 4). The putative ward clusters include a high proportion of suspected HAI, suggestive of linked transmission chains in the hospital.

**Figure 4.**
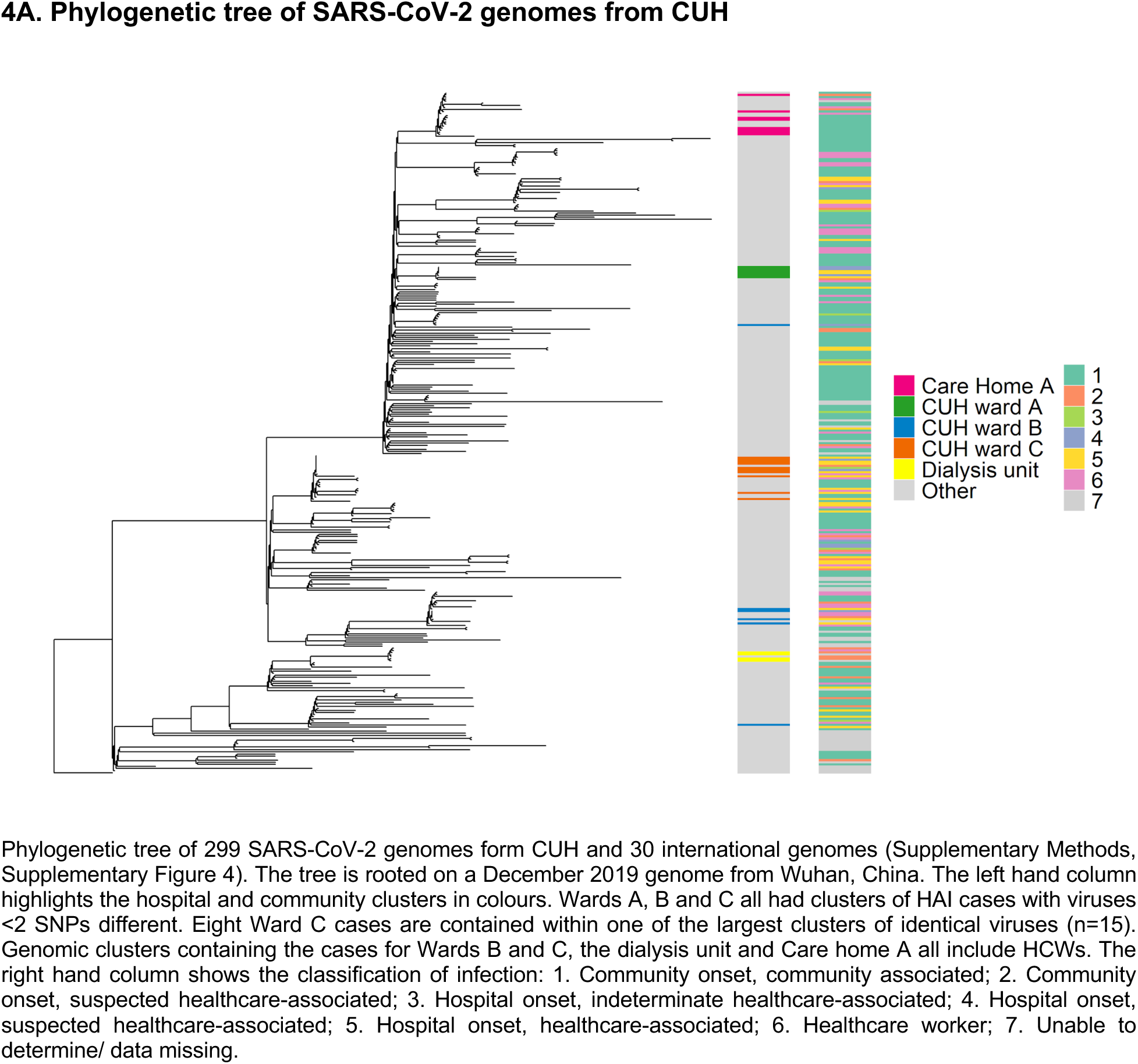
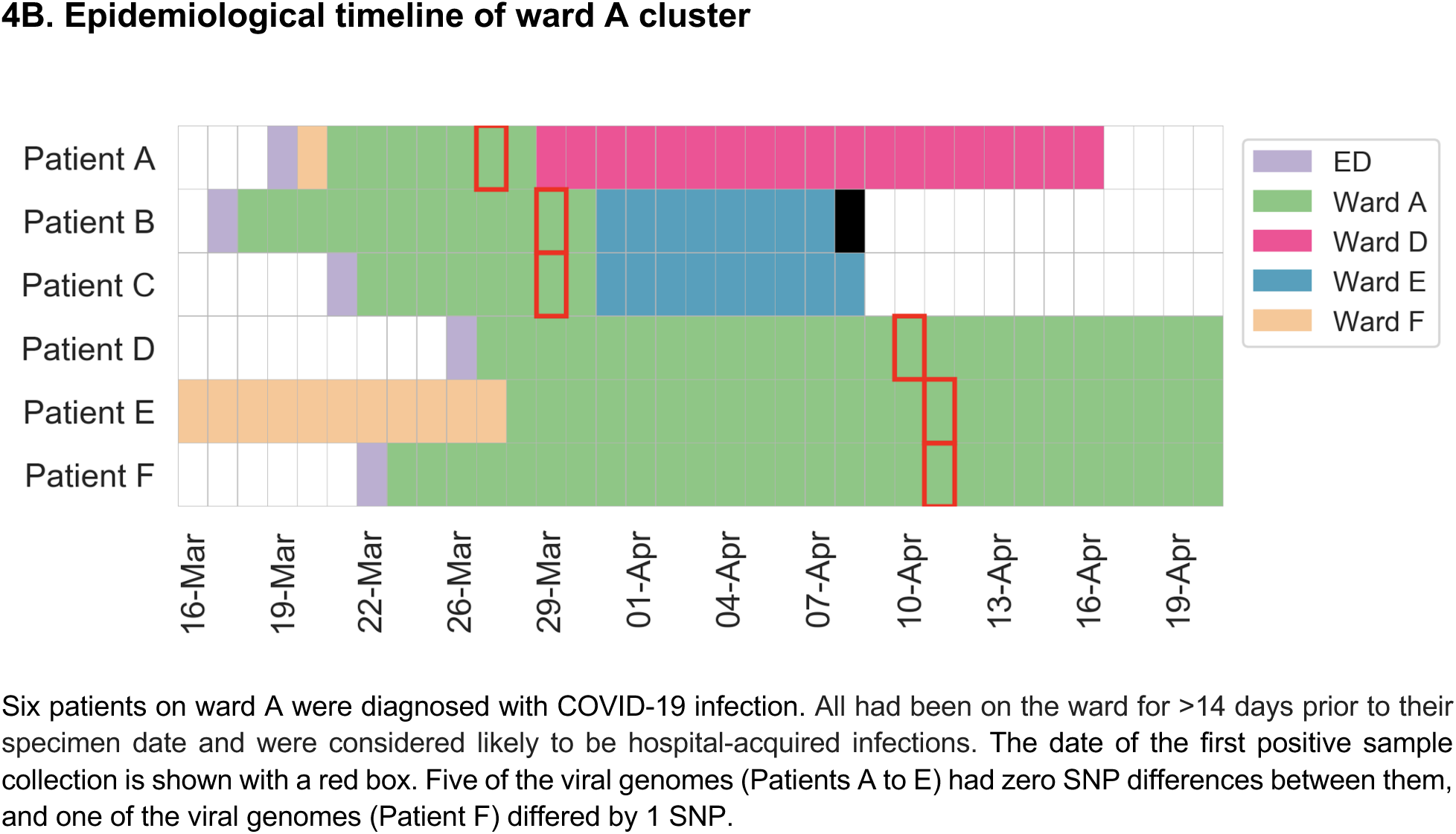
Phylogenetic tree and epidemiological timeline from CUH.

Due to its recent introduction into the human population, SARS-CoV-2 has low genetic diversity; here, a median of 8 single nucleotide polymorphisms (SNPs) separated any two samples at CUH (Supplementary Figure 5). This low sequence diversity makes interpretation of putative clusters challenging, as samples may be identical “by chance” rather than because of a recent connected transmission chain. To investigate genomic clustering further we adopted a combined genetic, clinical and epidemiological approach. Samples with zero SNP differences were identified and clusters named numerically by decreasing sample size (Supplementary Table 2).

Overall 114 genomes in 26 clusters shared at least one identical sequence. The two largest clusters each had 15 identical genomes. Patients’ medical records (including address, social setting, clinical details and ward movements) were reviewed for all putative genomic clusters to assess whether cases had plausible or likely linked recent transmission (Supplementary Methods). Of 114 cases from 26 putative clusters, 63 cases (55.3%) had strong epidemiological evidence to support recent transmission, 27 cases (23.7%) had intermediate evidence, and 24 (21.1%) no evidence of connected transmission. Clusters with strong evidence of linked recent transmission include cases where a connection was already suspected, such as groups of likely HAI seen on several CUH hospital wards (Figure 4), and clusters that were previously not recognised as being linked, such as a care home outbreak involving HCWs based in hospital and community settings. These are described further below.

### Application and impact of combined epidemiological and genomic analyses

Directed by clinical and infection control teams we established a process to conduct focused genomic and epidemiological analyses of suspected HAI cases (Supplementary Figure 6). These were discussed in weekly meetings with an accompanying written report submitted to the hospital. Anonymised examples are described here.

#### Hospital cluster 1

Six surgical patients on ward A (a non-COVID area) were diagnosed with COVID-19 infection (Figure 4B). All cases had been on the ward for >14 days prior to their specimen date and were classified as probable HAI. Genomic data were available for all cases; five differed by zero SNPs and one by a single SNP, consistent with recent ward-based transmission events. In response to this, universal mask use was introduced.

#### Hospital cluster 2

Four transplant patients on ward B (a non-COVID area) were diagnosed with COVID-19 infection between 3 and 20 April 2020 (Supplementary Figure 7A). A fifth patient, who had been recently discharged from the ward, presented to the ED with COVID-19 infection. Genomic analysis revealed that all 5 cases had identical genomes. Three HCWs were found to have identical genomes in the same cluster as the ward B cases; two had worked on ward B, one of whom had professional contact with the other HCW. These findings led to a review of infection control and PPE procedures for staff and patients in the transplant service.

#### Dialysis unit cluster

Renal dialysis patients are among the most vulnerable to COVID-19 (up to 19% mortality^20^), with challenging infection control arrangements as most units consist of large open rooms with no barriers between patients. Six patients with end-stage renal failure were diagnosed with COVID-19 between 1 and 20 April 2020, testing positive in several locations including ED and an acute admissions ward (Supplementary Figure 7B). Their viral genomes were identical, and epidemiological investigation revealed they dialysed at the same outpatient dialysis unit on the same days of the week. This suggests linked recent transmission of community-onset healthcare-associated infections. These findings led to a review of infection control procedures in dialysis patients and identified shared patient transportation as a possible risk factor. Genomics can also be used to “rule out” linked transmission. The renal ward (which shares patients with the outpatient dialysis unit) also had a group of COVID-19 cases at around the same time. However, the dialysis unit genomes belonged to lineage B.2 (relatively rare in EoE), whereas the renal ward genomes were the common B.1 lineage, making it very unlikely that infections between the two patient groups were related.

#### Community cluster

Fifteen patients were admitted to CUH with COVID-19 between 7 and 21 April 2020 with genetically identical viruses. Eight patients were residents at a community care home (care home A). A review of the medical records revealed that another patient in this genetic cluster worked in care home A and one was a retired nurse who worked in an unknown care home. Two cases were paramedics and two were nurses (who worked in different wards at CUH but lived with paramedics). The final case did not have any discernible epidemiological links with the others. In summary, this investigation revealed a cluster of cases with evidence of linked transmission coming from the same care home with potential links between HCW and the care home, none of which had been detected by clinicians or infection control.

The information from these combined epidemiological / genomic investigations was fed back to the clinical, infection control, and hospital management teams. This triggered further investigations of patient isolation, ward cleaning procedures, use of personal protective equipment (PPE) and staff social distancing behaviour. HCAI were also assessed in relation to potential harm caused to patients and recorded in hospital’s patient safety reporting system for follow-up and further action.

## Discussion

The value of real-time viral genome sequencing has been demonstrated in previous epidemics ^21-26^ We sought to embed genomic surveillance as part of an active SARS-CoV-2 infection control process in a large UK hospital. A rapid sequencing workflow was established on 23 March 2020 using multiplex PCR-based nanopore sequencing, which has been proven to be effective in a wide range of clinical samples and viral loads ^27^. We aimed to sequence all available positive samples from CUH and a selection from each of the EoE regional hospitals submitted to the diagnostic microbiology laboratory, linking with clinical metadata pulled from the hospital electronic patient records system. We also included 44 samples collected as part of a local HCW screening programme (Rivett et al, unpublished). In 5 weeks, over 1,000 SARS-CoV-2 genomes had been sequenced including the majority of CUH samples from this phase of the epidemic. We applied this system to investigate nosocomial and HCW COVID-19 infections at CUH, integrating genomics with epidemiological and clinical data.

We examined the diversity in SARS-CoV-2 at CUH and found that overall genetic diversity was low and reflected the pattern seen in the EoE as a whole, with most viruses belonging to the B.1 lineage. We identified a median of 8 (range 0 to 24) SNP differences between viruses, and 4.5% of pairwise comparisons between CUH genomes had 0 to 1 SNP differences. Given the virus’s mutation rate and infectious timeframe, cases may share linked recent transmission if there are fewer than ~2 SNP differences. However, unravelling potential transmission networks requires rapid integration with in-depth epidemiological data.

We investigated clusters of patients and HCW cases at CUH in response to queries from the clinical and infection control teams. Using ward location data for patients and HCWs we performed epidemiological analyses to determine if there had been ward-based contact. We compared the genomes of patients and HCWs in the suspected clusters with those of other patients at CUH and in EoE hospitals to examine relatedness. This approach enabled us to confirm or refute evidence for transmission between patients and / or HCWs within these clusters. Genomic analysis also enabled us to identify cryptic transmission i.e. additional cases that were not initially suspected to be linked to the original ward or HCW clusters. We were also able to use genomic data to refute suspected transmission events (e.g. different lineages between two isolates that were epidemiologically linked), also important as a mechanism to monitor infection control measures. These cases illustrate the power of combining rapid genomic and epidemiological analyses in near real time. In contrast to previous studies^28–31^, we reported results of our investigations to the clinical, infection control and management teams on a weekly basis, thus enabling them to respond to this information and act accordingly within the timeframe of ongoing ward outbreaks. The genomic data informed reviews of patient placement and isolation procedures, assessment of PPE use and staff break arrangements, supporting us to better focus efforts at a time of unprecedented demand on infection control teams. Finally, these analyses are currently being used to inform existing patient safety review processes within our hospital, including investigations related to hospital-onset COVID-19 where the patient has come to harm.

Our study highlights the importance of understanding SARS-CoV-2 transmission within healthcare settings in managing the pandemic. The possible transmission networks that we have identified were complex, involving patients and HCW in both community and hospital settings. However, we have also identified potential transmission networks in other environments including care homes, outpatient units and ambulance services, that have been poorly studied. Although there are strong epidemiological and genomic associations between cases, the mechanism and direction of transmissions remain unclear. The role of asymptomatic intermediates, fomites (including PPE) and the environment are not well understood and require further investigation. During the time frame of this study a number of infection control interventions were implemented across the hospital, as well as national public health measures to reduce community spread. Understanding the interaction between such interventions and nosocomial transmission are complex (especially in the context of the comparably long incubation period for SARS-CoV-2 relative to other respiratory viruses), but critical in enabling healthcare providers to safely deliver existing services in the context of a pandemic.

We acknowledge several limitations to our study. Firstly, we were unable to sequence all of the genomes from samples that were collected during the study period. We may therefore have missed the opportunity to investigate all potential transmission events. Furthermore, we only had main ward location data for most of the HCW and could have overlooked potential epidemiological links with patients with whom they had contact on other wards. Similarly, we were unable to track HCW movements outside their main ward location to identify potential contact with patients on other wards or HCW within shared communal areas. Finally, due to the recent introduction of SARS-CoV-2 into the human population, highly similar genomes are not sufficient evidence to infer a recent transmission, in the absence of confirmatory epidemiological data. However, our experience indicates that further investigation of genomic clusters with highly similar genomes can uncover previously unknown epidemiological links. Furthermore, we were able to use rapidly generated genomic data to investigate HCAI within the hospital setting. The next initiative is to expand this platform into a more detailed analysis of infections in HCW and in community settings, such as residential homes. As the practical challenges associated with implementing real-time genome sequencing during epidemics are overcome, unlocking the real power of genomic epidemiology will require its integration with clinical and public health systems to support decision-making on local, national and international scales.

## Data Availability

Genome sequence data from this manuscript are available from GISAD https://www.gisaid.org/

## Acknowledgments

We acknowledge the assistance of the laboratory staff of CMPHL for processing the diagnostic samples and the clinical teams (infectious diseases, microbiology, virology, infection control) at CUH for their assistance with the investigation of healthcare-associated infections. We are grateful to Richard Smith (CUH Patient Safety Lead), Lucy Rivett, Dominic Sparkes, Nick K. Jones and Matthew Routledge (for providing some HCW data), and Anthony Underwood (Centre for Genomic Pathogen Surveillance) for helpful discussions and advice.

## Supplementary Methods

### SARS-CoV-2 molecular testing

Nucleic acid extraction was undertaken using the NUCLISENS easyMAG platform (Biomerieux, Marcy L-Etoile), in accordance with manufacturers’ instructions. Nucleic acids were extracted from 500μL of sample, with a dilution of MS2 bacteriophage added pre-extraction to act as an internal extraction and inhibition control. The presence of SARS-CoV-2 was assessed using an in-house generated and validated one-step RT q-PCR assay that detects a 222 base-pair region of the SARS-CoV-2 RdRp genes, along with an MS2 bacteriophage internal extraction control. The RdRp gene was detected using the RdRp For primer (ATGGGTTGGGATTATCCTAAATGTGA) and the RdRp Rev primer (AGCAGTTGTGGCATCTCCTGATGAG) with a FAM labelled MGB RdRp Probe 3 (ATGCTTAGAATTATGGCCTCAC). The internal extraction control was detected using the MS2 For primer (TGGCACTACCCCTCTCCGTATTCACG), the MS2 Rev primer (GTACGGGCGACCCCACGATGAC) and a ROX-BHQ2 labelled MS2 probe (CACATCGATAGATCAAGGTGCCTACAAGC). Amplification reactions and detection of PCR products were performed using the Rotorgene™ PCR instrument. A typical reaction contained 400nM of For and Rev primers for the RdRp genes and 200nM of the the MS2 internal control For and Rev primer pair, along with 120nM of the RdRpand MS2 probes. Reactions typically contained 25% extracted nucleic acid and were cycled through the following conditions 25°C 2min, 50°C 15min, 95°C 2min followed by 45 cycles of 95°C and 60°C. Samples that generated a Ct value ≤36 were considered positive. Samples and negative control (molecular grade water) were individually spiked with MS2 bacteriophage internal control (4600 pfu per extraction) prior to nucleic acid extraction to identify any inhibitors or extraction issues. Positive control material, BetaCoV/England/02/2020, was obtained from PHE Colindale and was essentially purified virus RNA diluted down to give a cycle threshold value of 26-28. Negative controls included extracted molecular grade water.

### Sample and data collection

Sample collection, metadata curation and linkage to sequencing IDs involved coordination between multiple teams and critically depended on good relations between the sequencing and diagnostic laboratory staff (Supplementary Figure 1). Each day samples with positive SARS-CoV-2 PCR results from the last 24 to 72 hours were identified from the hospital information system and clinical metadata was extracted, formatted and integrated into a master metadata file. A 15 microlitre aliquot of the RNA extract of all CUH samples and a random selection of EoE samples were collected from the diagnostic microbiology laboratory for local sequencing. These were assigned COG-UK sequencing codes which were integrated back into the master metadata file. For samples that were not sequenced locally, the remaining RNA extract was collected from the diagnostic microbiology laboratory and sent to the Wellcome Sanger Institute for sequencing as part of the COG-UK consortium. Each week, clinical metadata and sequencing data were combined and formatted for upload to the MRC CLIMB system. Data manipulations were performed in R (v 3.6.2) using the *tidyverse* packages (v 1.3.0) installed onto computers within the Trust network.

### Sequencing details

Samples were sequenced using Nanopore technology following the ARTICnetwork V3 protocol (https://dx.doi.org/10.17504/protocols.io.bbmuik6w) and assembled using the ARTICnetwork assembly pipeline (https://artic.network/ncov-2019/ncov2019-bioinformatics-sop.html). Median genome depth of coverage was 6,612x across all 747 genomes. 14 samples in our dataset were also sequenced with Illumina technology at the Wellcome Sanger Institute as part of COG-UK. There was 100% concordance in called nucleotides between sample pairs. Four genomes differed because of base pairs called in the Illumina data that were missing in the Nanopore sequences. The accession numbers of the samples included in this study are available in Supplementary Table 4.

### Bioinformatic analysis

Consensus fasta sequence quality control cutoffs were: size >29Kb, N count <2990 (~10%). After QC filtering, de-duplication and matching with metadata, the first sample set analysed comprised 197 genomes collected up to 10th April 2020; set 2 had 444 genomes up to 15th April, and set 3 (presented here) had 747 genomes up to 24th April. 30 reference genomes were added to the sample sets downloaded from GISAID (https://www.gisaid.org/; Supplementary Table 3). The reference genomes were chosen to represent the major branches of the global phylogenetic tree as visualised in Nextstrain (https://nextstrain.org/) to provide broader context, including a sample from December 2019 collected in Wuhan, China, used to root the tree. Multiple sequence alignment was performed using MAFFT (v 7.458) with default settings, command:

/PATH/mafft” --retree 2 --inputorder “multi_fasta.fasta” > “aligned_multi_fasta The alignment was manually inspected using AliView. Maximum likelihood trees were produced using IQ-TREE software^15^ for all samples passing QC filters and the subset of samples from CUH (n=299 for this dataset). Initial tests with the ModelFinder Plus option^32^, which selects the optimal nucleotide substitution model out of over 200 options (http://www.iqtree.org/doc/Substitution-Models), consistently identified GTR+F+I as the best model. Therefore from 24th April (including analysis presented here) we specified GTR+F+I.

Command using ModelFinder Plus:

/PATH/iqtree -s aligned_filtered_multi_fasta -m MFP

Command with GTR+F+I model specified:

/PATH/iqtree -s aligned_filtered_multi_fasta -m GTR+F+I

Trees were manually inspected in FigTree, rooted on the 2019 Wuhan sample (EPI ISL 402123), ordered by descending node and exported as Newick files. Trees were visualised in online software Microreact^16^ in a private account to explore relationships between wards and clinico-epidemiological questions for our weekly reports. Further visualisations were produced in R using the packages *Ape*^33^ (v 5.3) and ggtree^17^ (v 2.0.4).

A SNP difference matrix was produced from the multiple sequence alignment using the *snp-dists* package (v 0.7.0; https://github.com/tseemann/snp-dists; installed into a conda environment), command:

snp-dists-c aligned_filtered_multi_fasta.aln > snp_dist_matrix.csv

The matrix was exported as .csv and manipulated in R using the *Matrix* and *tidyverse* packages for ward and pairwise SNP comparisons and plotted using the *ggplot2* (v 3.3.0) package. A heatmap was produced in python using the seaborn (v 0.10.0) clustermap function. To identify clusters with zero SNP differences we used the scipy.cluster.hierarchy functions *linkage* and *fcluster* setting a maximum distance of zero (scipy v 1.4.1). Clusters were named in descending size order and linked with sample metadata and lineage data.

Investigation of genetic clusters with zero SNP differences

Patient records from each case within a putative genomic cluster were manually reviewed in detail by authors BW, WLH and MET and assigned a score of 1 (strong evidence supporting a recent linked transmission chain, e.g. patients co-located on the same ward becoming positive within the incubation period of the virus), 2 (a plausible transmission chain is present e.g. patients becoming positive while located in nearby wards within the hospital but who did not appear to be in direct contact), and 3 (no evidence of any transmission connections between cases).

### Epidemiological analysis methods

#### Timeline plotting

Space time relationships between patients were plotted using patient specific time lines by exporting the bed and ward admission dates, dates of transfer, dates of discharge or date of death obtained from hospital information system (EPIC Systems Corporation, Verona, USA) and importing them into a cloud-based timeline plotter application (Cluster Track, Camart Ltd, Cambridge available at Clusterack.com). Earliest positive specimen date for COVID19 was obtained from the laboratory records and date of onset of symptoms from the clinical records and uploaded.

The application aided visualisation of ward and time relations by assigning unique colours to wards and then ward presence by date along the x axis shown in days, such that a solid timeline bar by colour and by date permitted the visualisation of the location of each patient over time. The positive specimen date for COVID 19, genomic cluster, death and discharge were each overlaid on the patient timeline using standard visual representation built into the application. Visualisation was aided by using the sort command within the application on admission date, earliest positive specimen date, or first ward to which admitted. Separate plots of subset of the total cases were created to provide clearer visualisation when needed

#### Ward time and genomic cluster plots

A clustering and network analysis function was used in the Cluster track application in which an algorithm links patients with admission days to the same ward on the same date and displays a network diagram to indicate these overlapping cases.

More advanced space time clustering was undertaken by exporting these timeline data sets into an SQL database running a more advanced clustering algorithm in which time parameters were set for the presumed susceptible, infectious and non-infectious/ recovered intervals counting from the earliest positive specimen date. The algorithm identified and linked cases in which two or more patients had an overlap on the same ward of the time interval of infectiousness of an earlier case with the interval of susceptibility in a later case or cases. Links continue to be made until no further overlaps of the infectious interval in an earlier case occurred with the interval of susceptibility in a later case on the same ward: this ended the space time cluster.

The cluster diagrams of the space time clustered cases were reviewed. Cases within the same space time cluster that belonged to the same genomic cluster were deemed to be supportive of a recent transmission event.

**Supplementary Figure 1.**
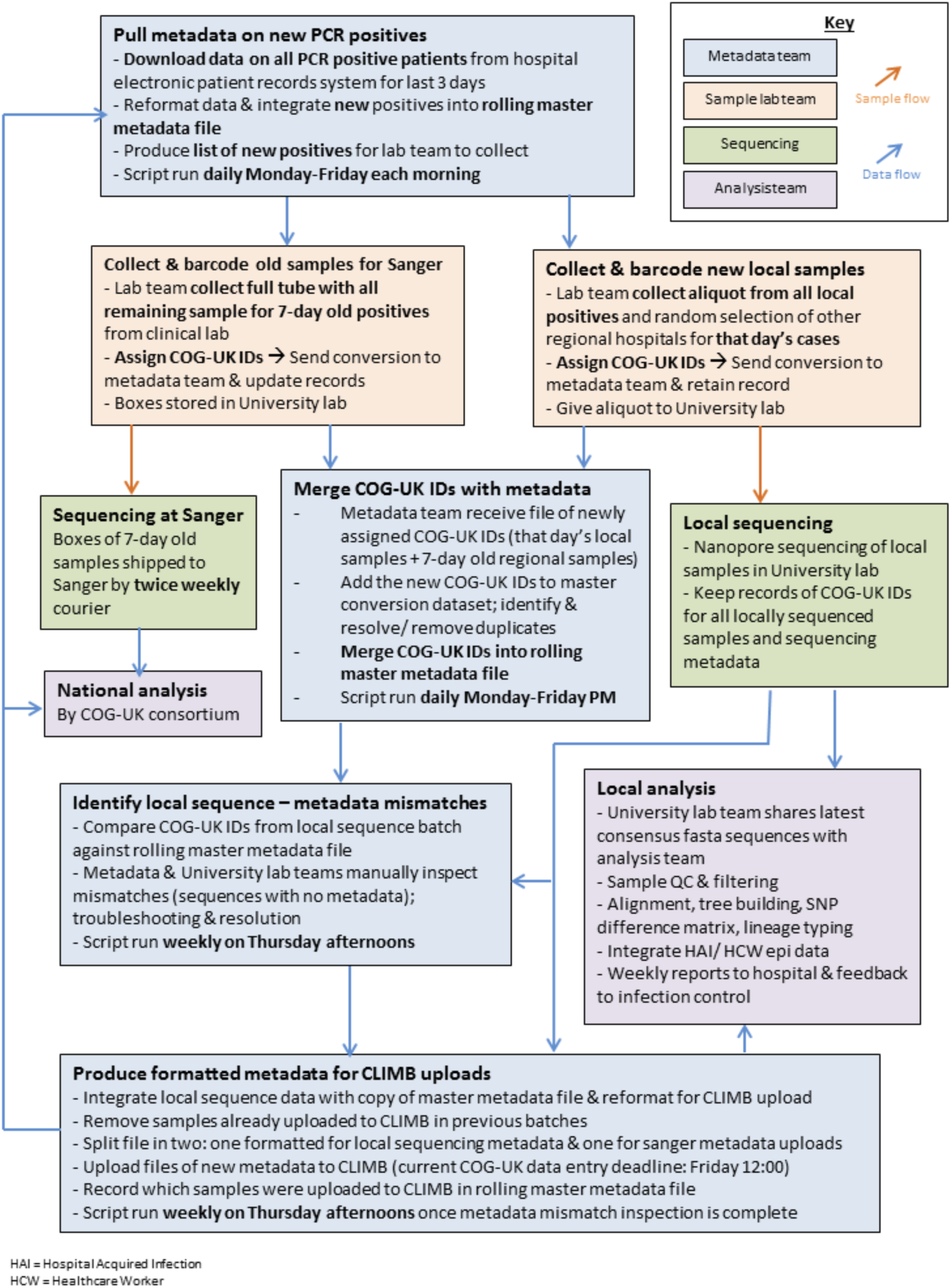
Data and Sample Processing at CUH.

**Supplementary Figure 2.**
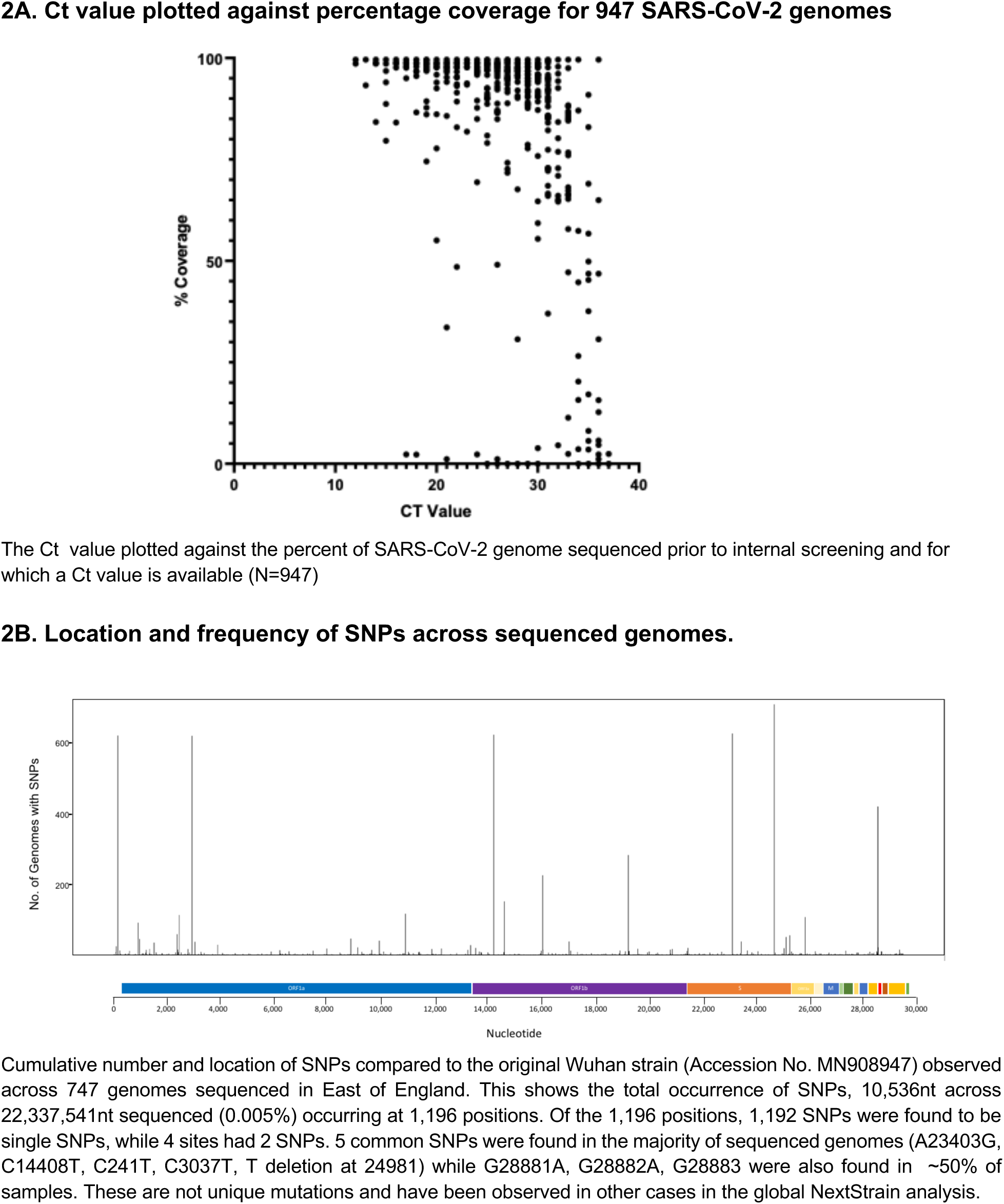

**Supplementary Figure 3.**
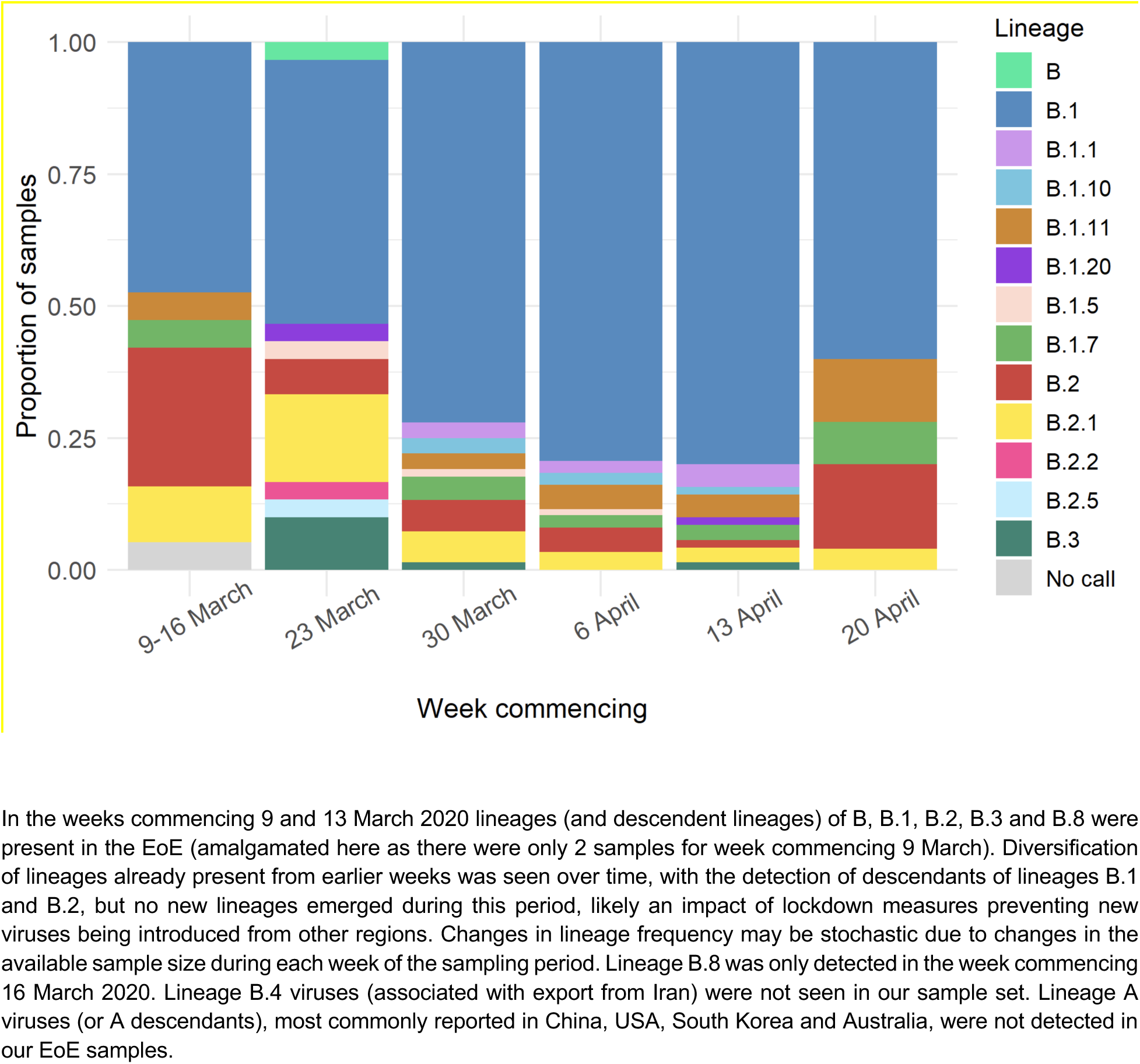
SARS-CoV-2 lineages over time.

**Supplementary Figure 4.**
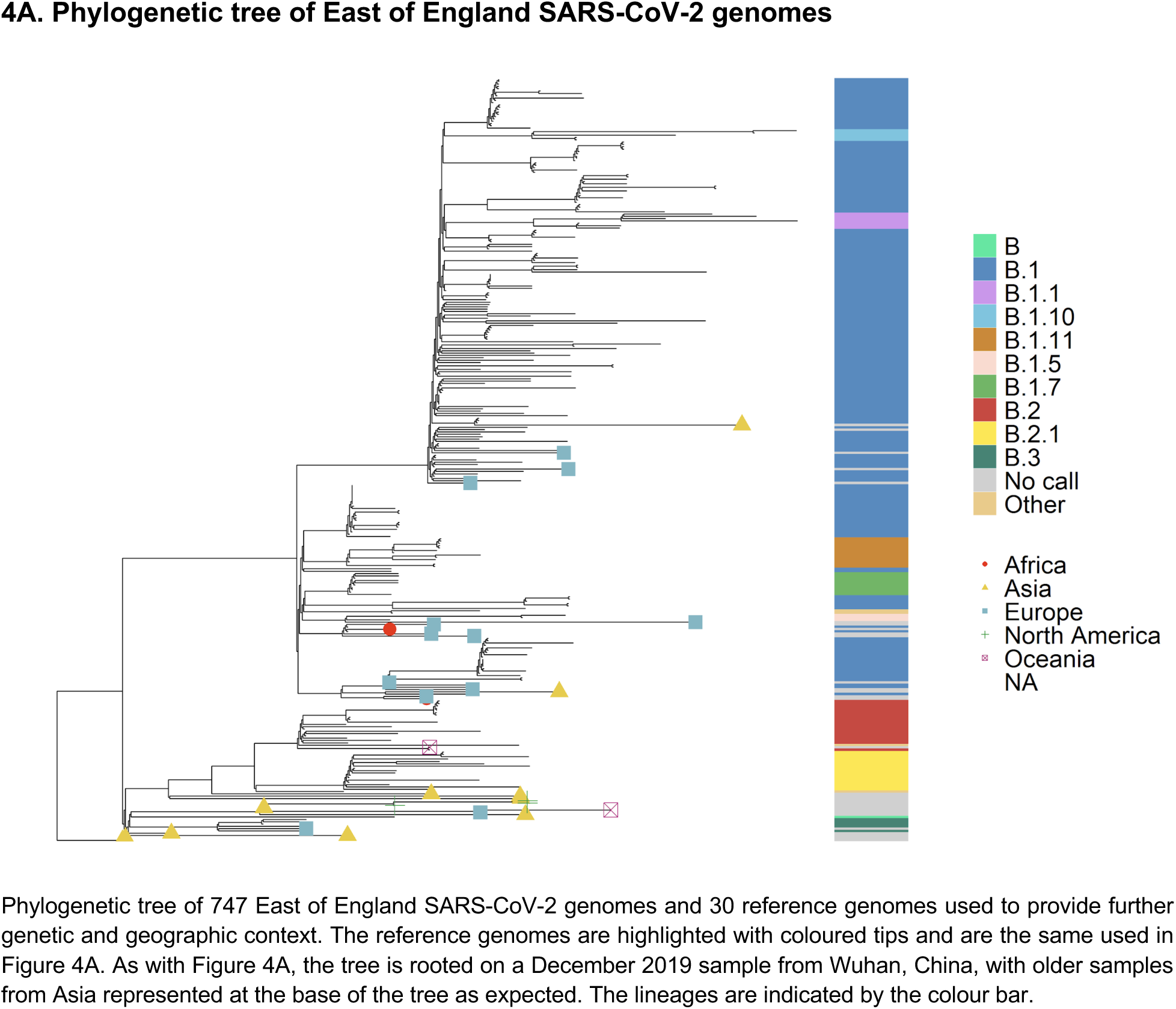
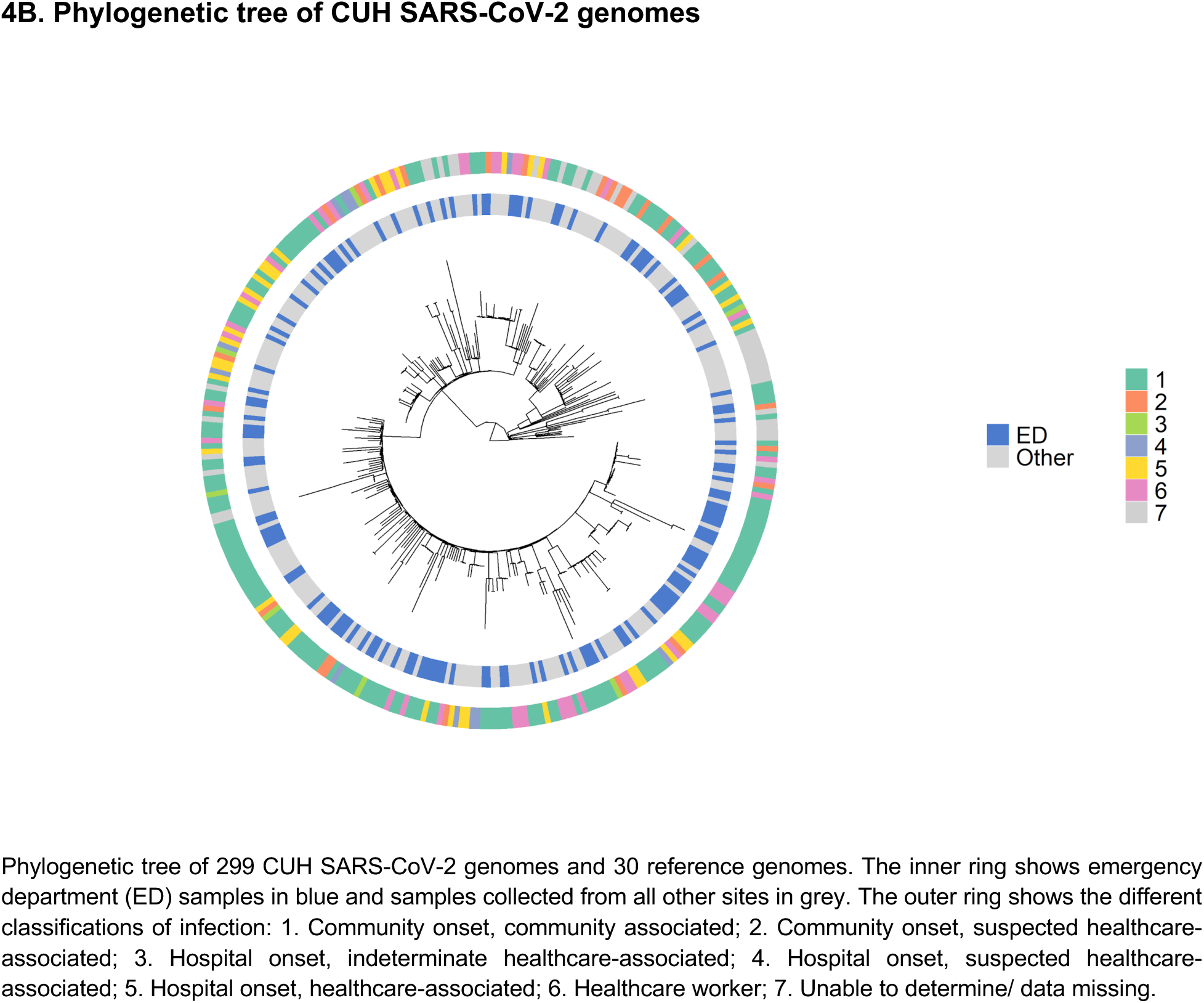
Phylogenetic trees of East of England and CUH genomes.

**Supplementary Figure 5.**
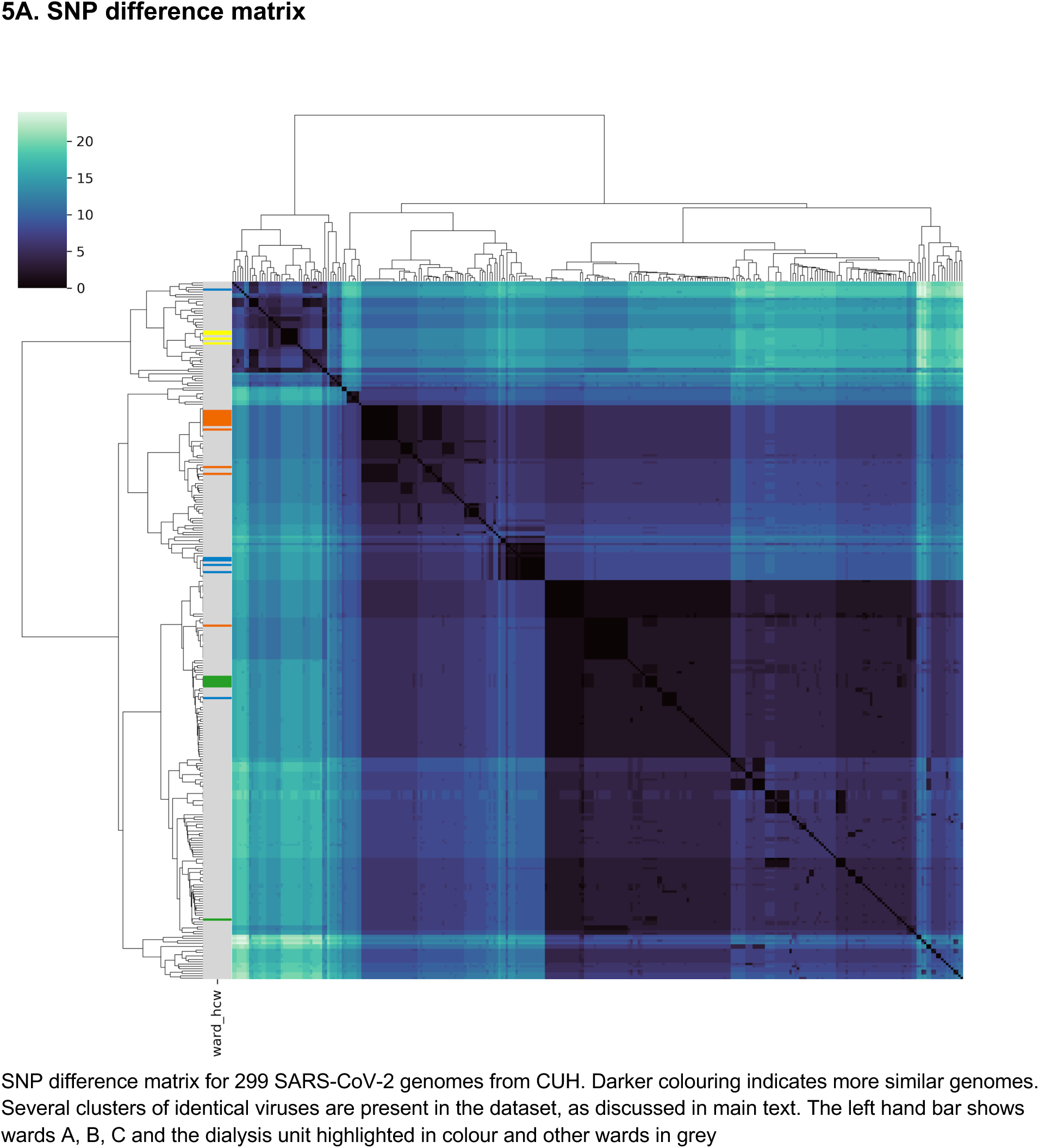
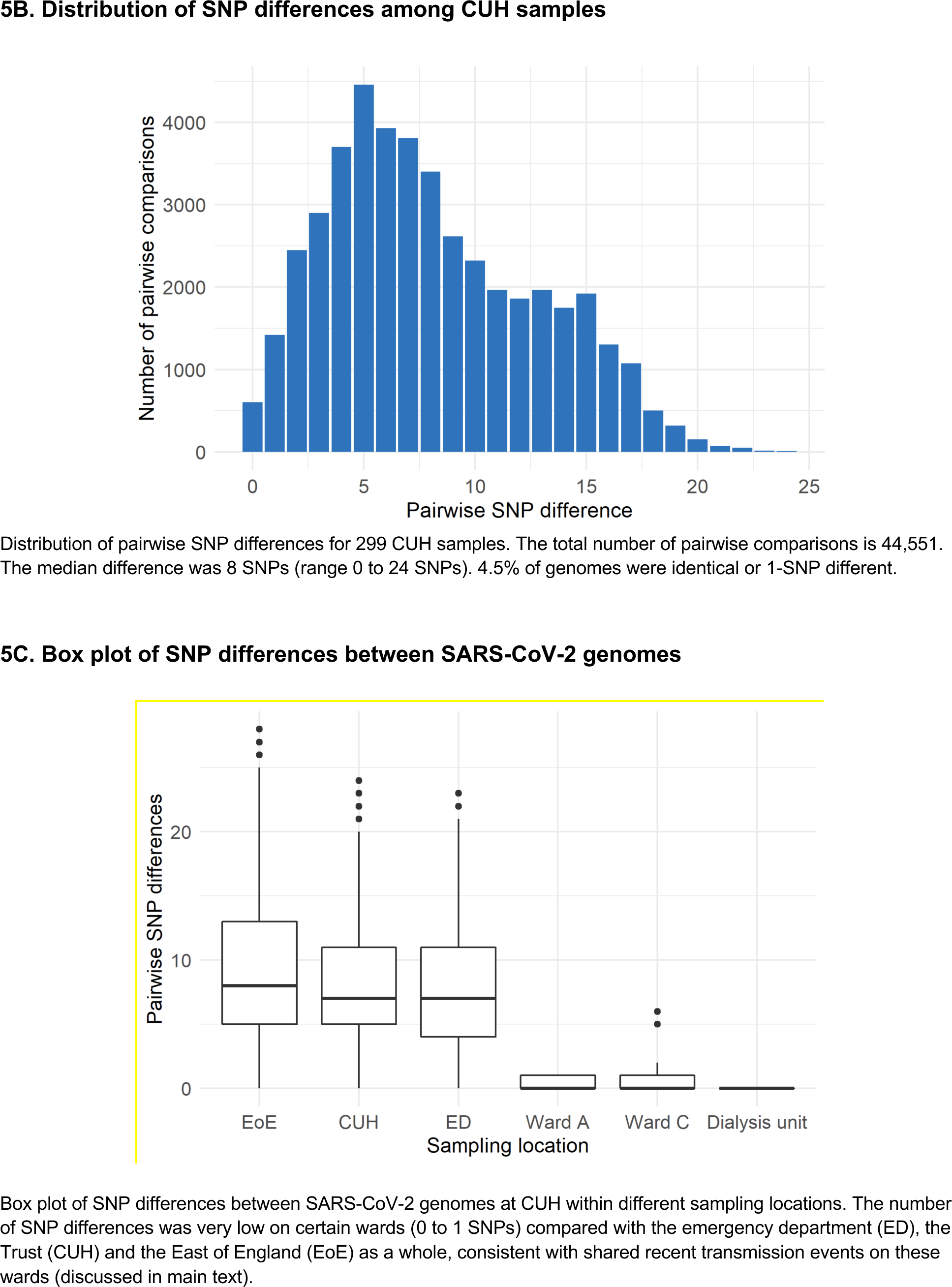
SNP difference matrix, distribution and box plots.

**Supplementary Figure 6.**
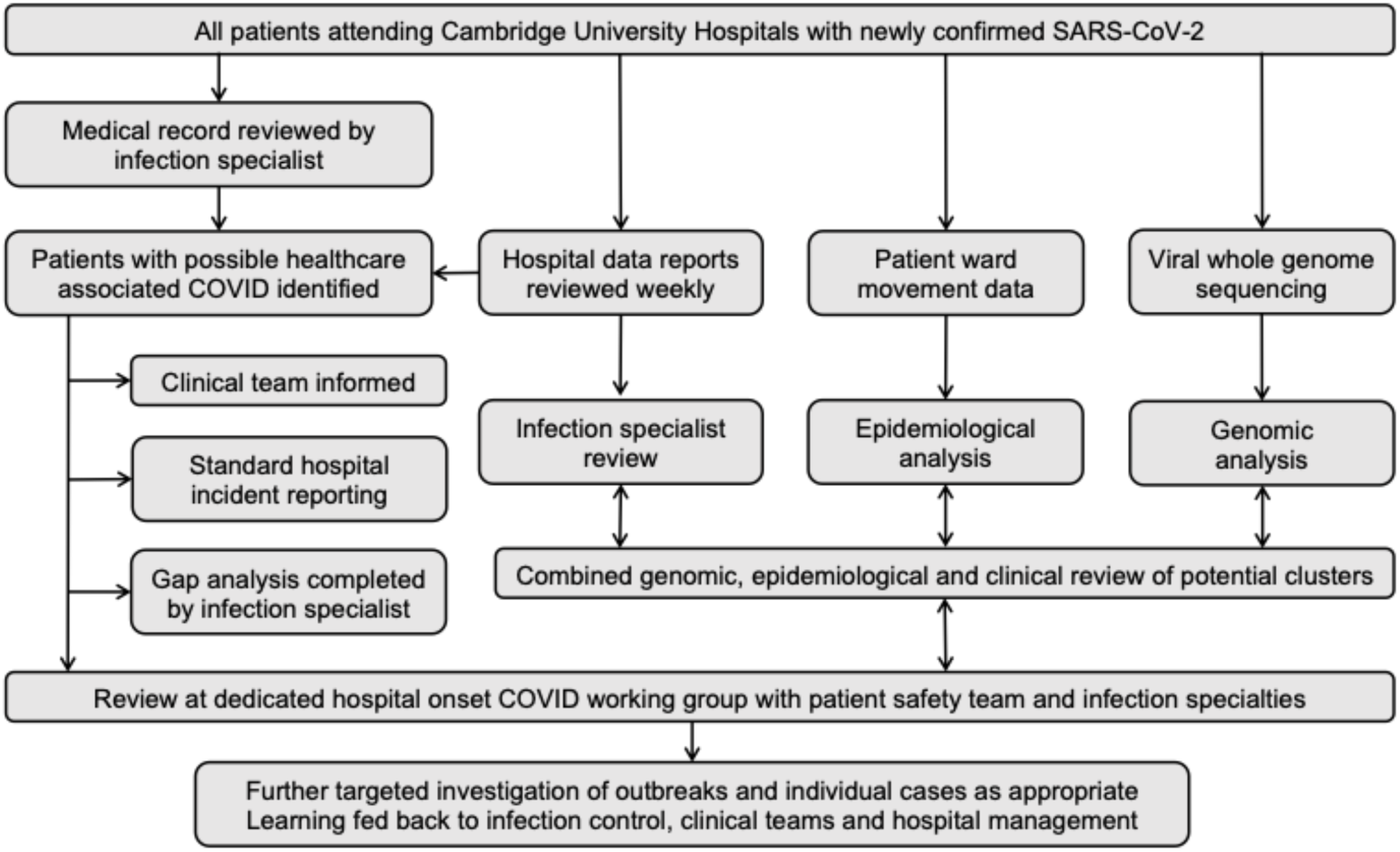
Process for investigating healthcare associated COVID-19 infections at Cambridge University Hospitals NHS Foundation Trust.

**Supplementary Figure 7.**
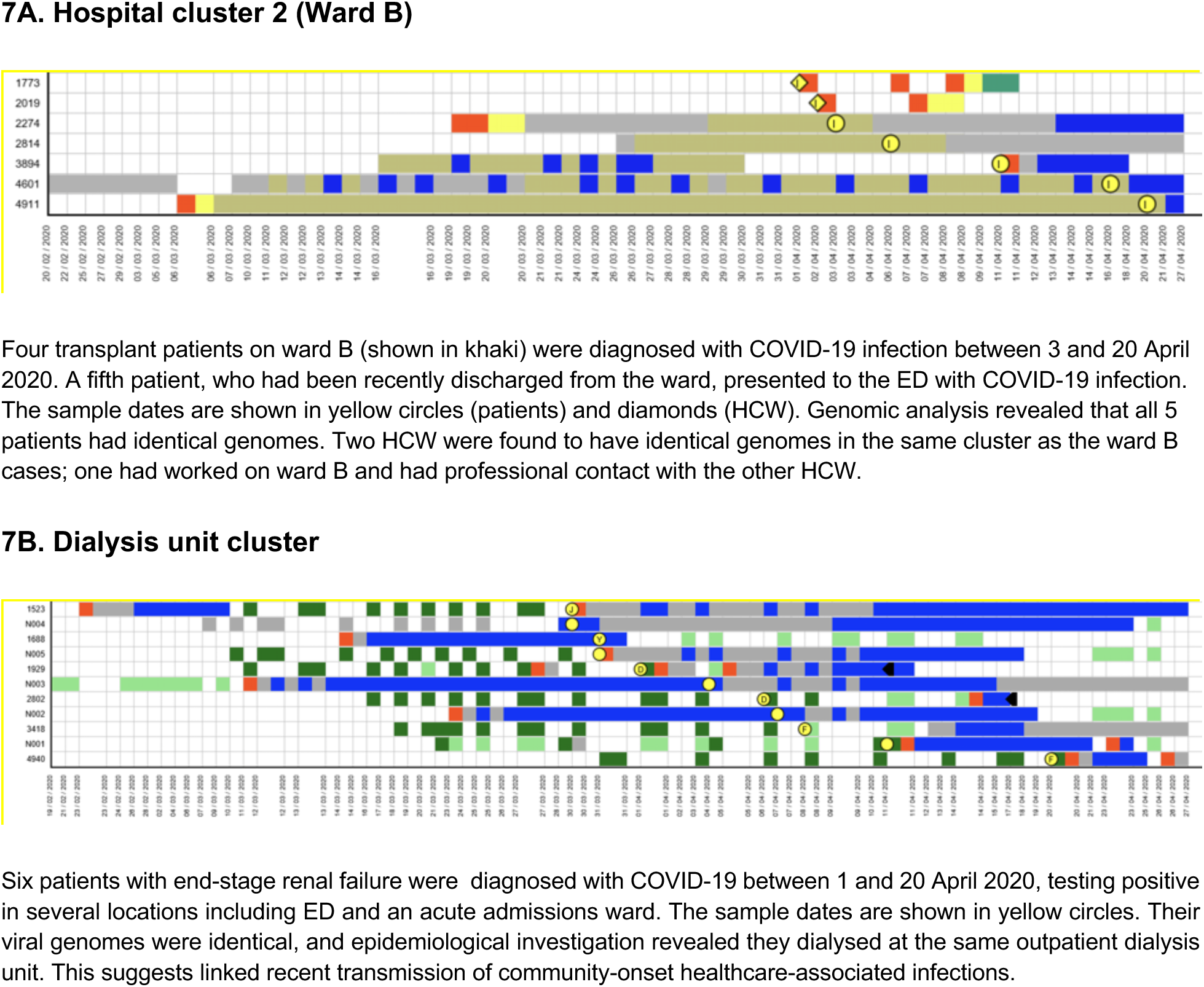
Epidemiological timelines of hospital clusters.

**Supplementary Table 1.** CUH COVID-19 infections and sequence data availability.

| **Classification of infection** | **No.** | **No. with available sequence (%)** |
| --- | --- | --- |
| Community onset, community associated | 261 | 174 (66.7%) |
| Community onset, suspected healthcare-associated | 40 | 27 (67.5%) |
| Hospital onset, indeterminate healthcare-associated | 11 | 7 (63.6%) |
| Hospital onset, suspected healthcare-associated | 13 | 11 (84.6%) |
| Hospital onset, healthcare-associated | 39 | 36 (92.3%) |
| Healthcare worker | 9 | 6 (66.7%) |
| Unknown | 1 | 1 (100%) |
| Total | 374 | 262 (70.1%) |

**Supplementary Table 2.**
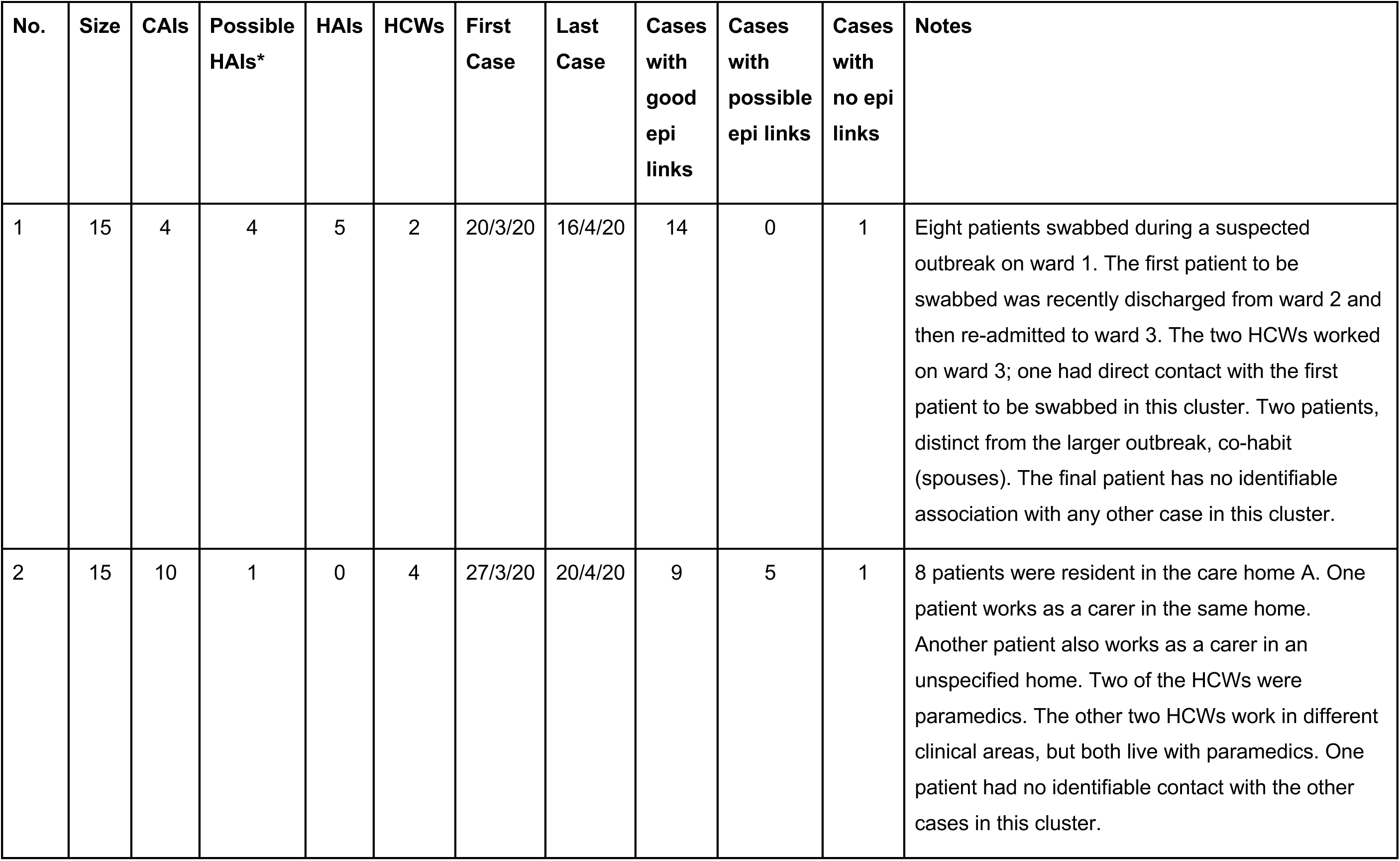
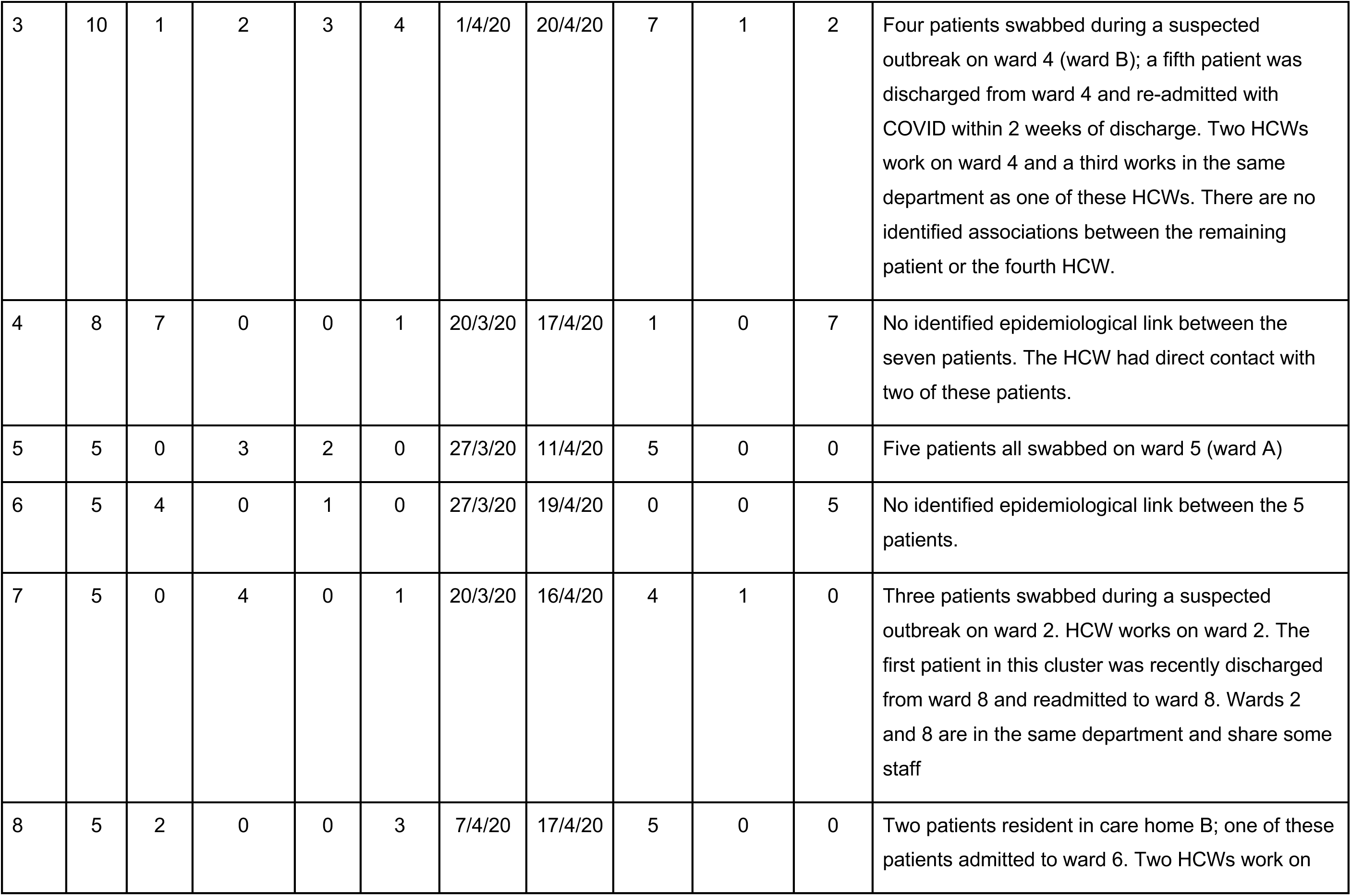
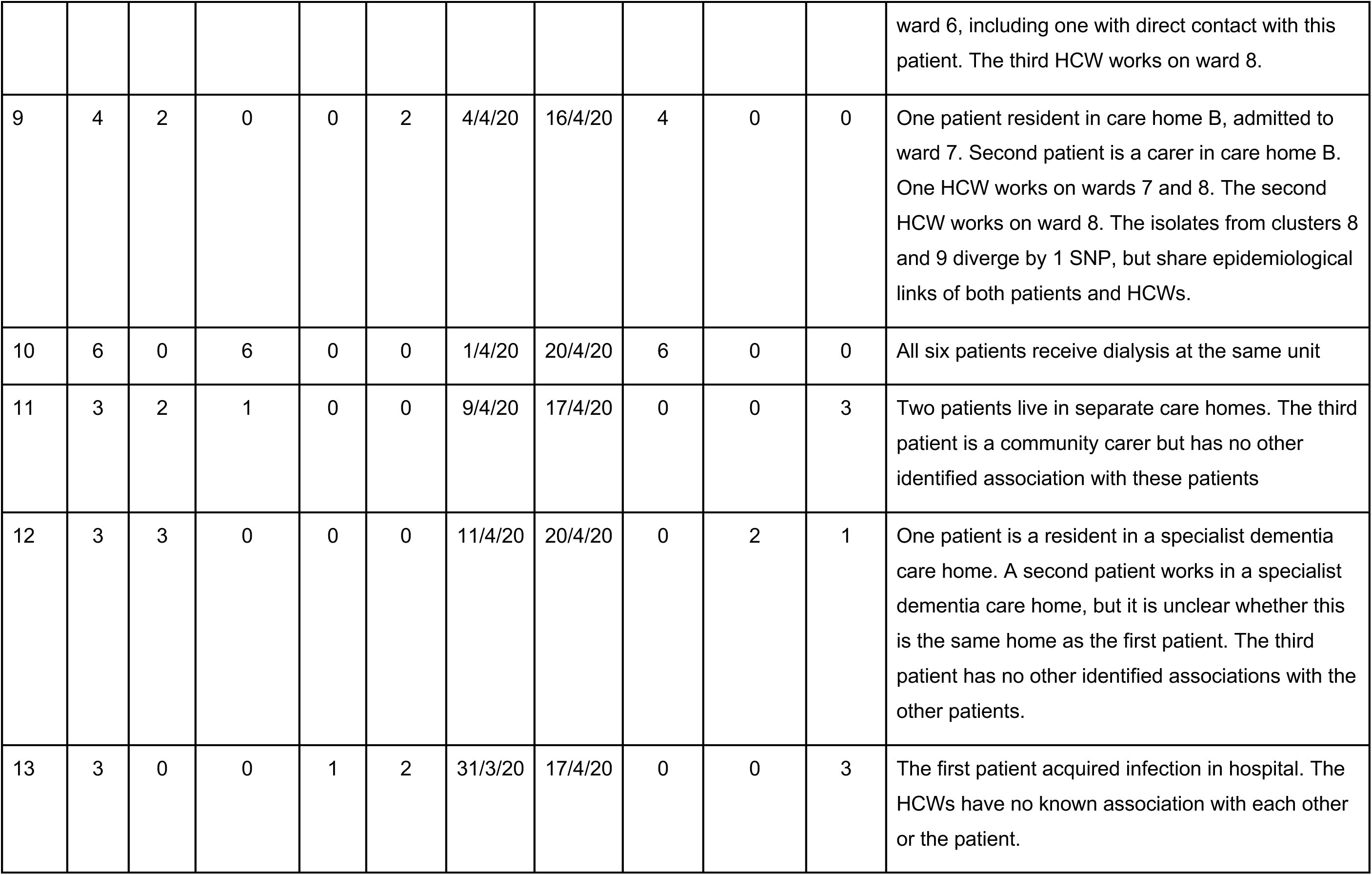
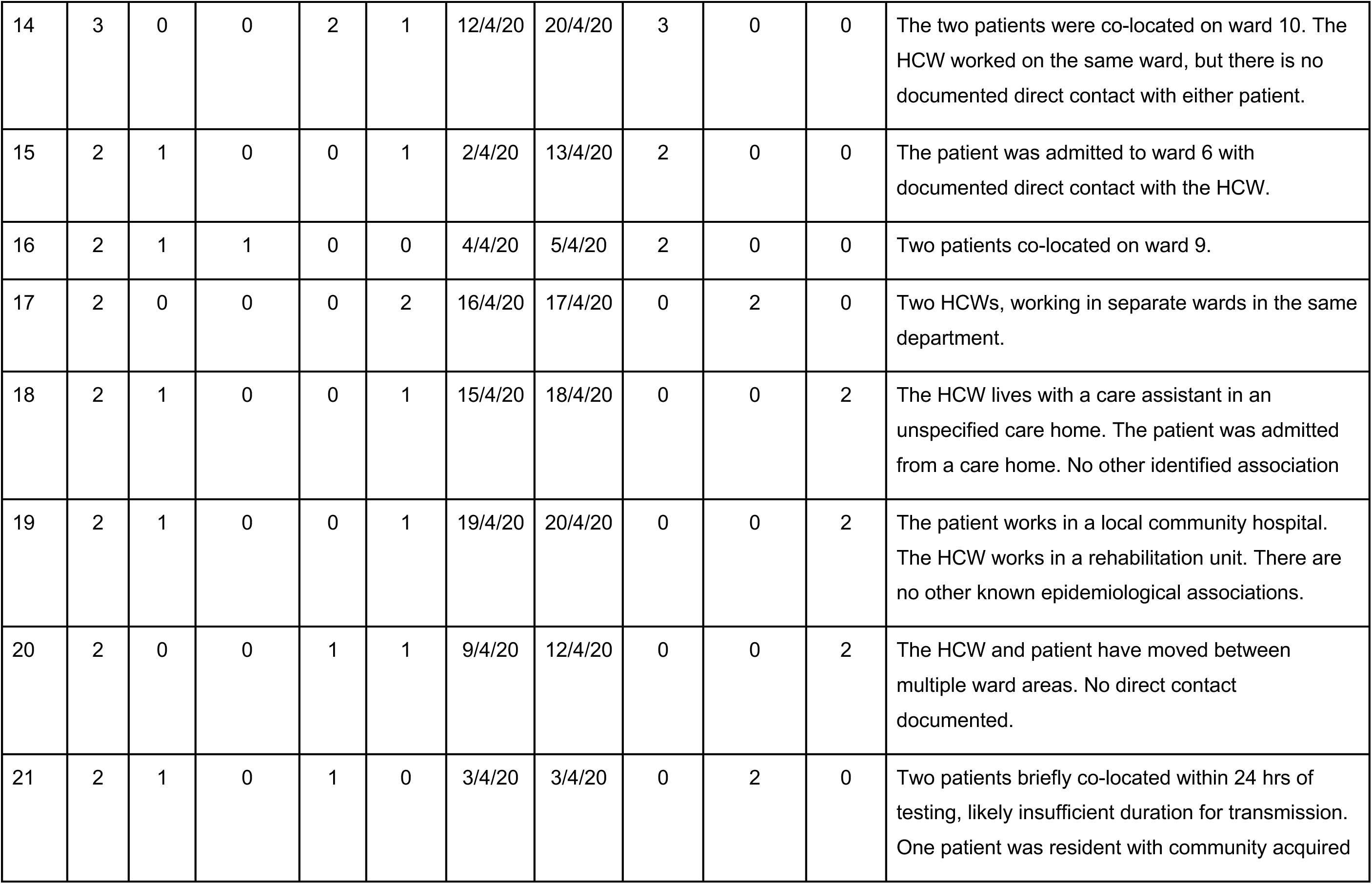
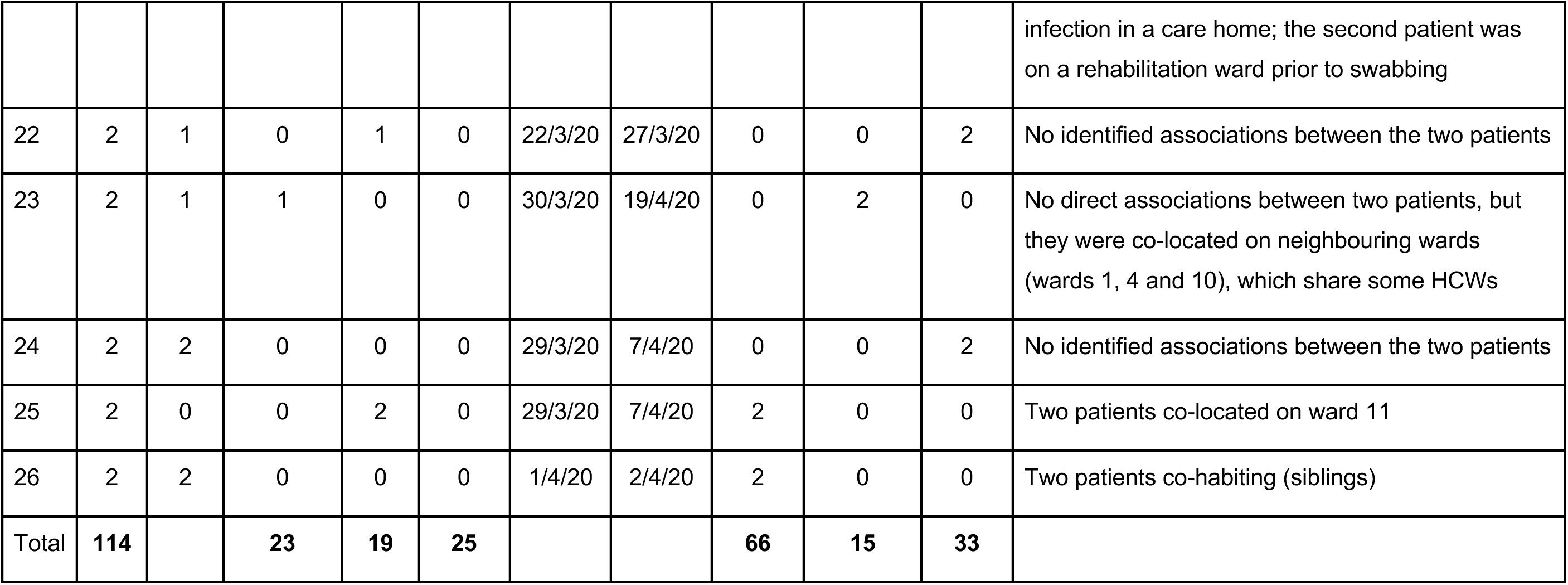
Transmission cluster table.

**Supplementary Table 3.**
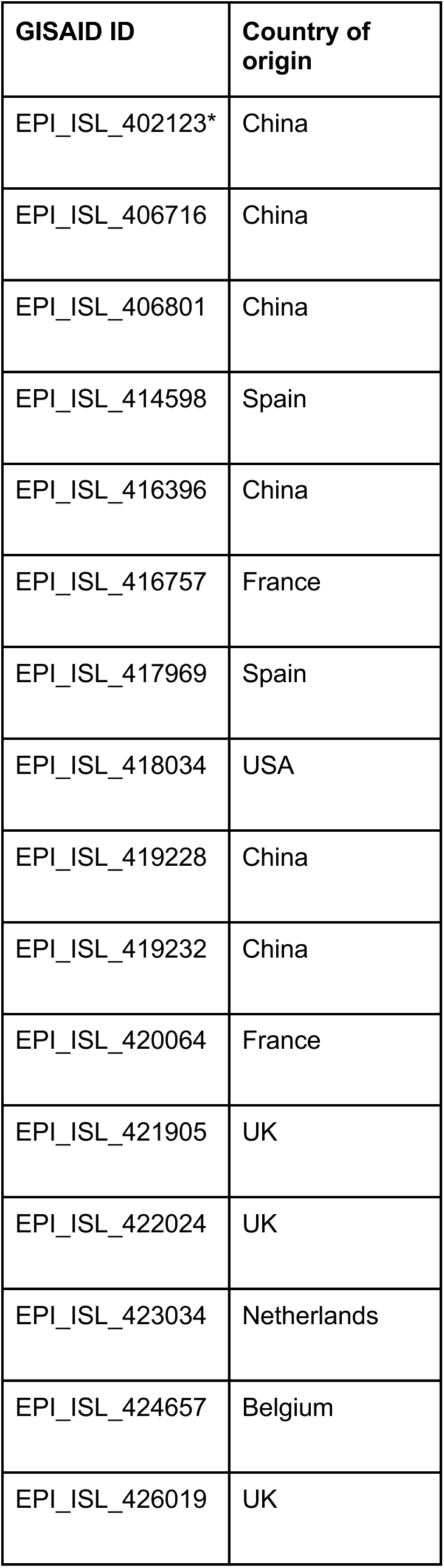
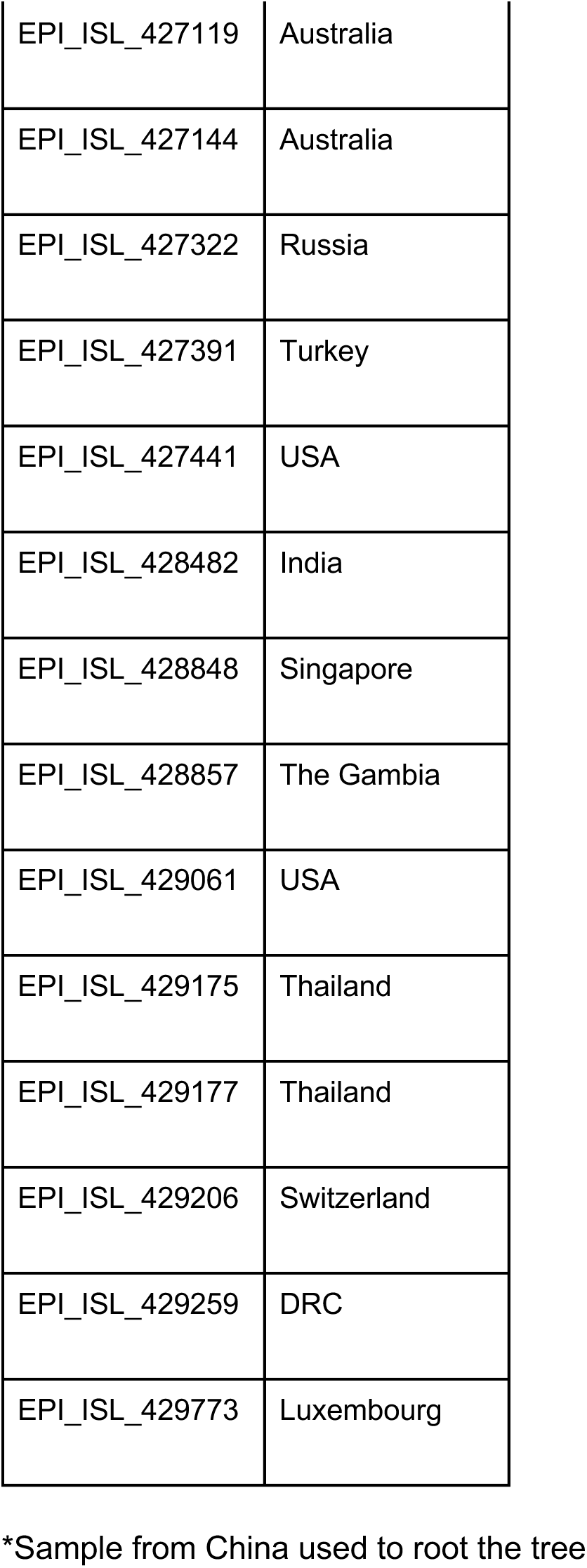
GISAID genomes included in phylogenetic tree.

**Supplementary Table 4.**
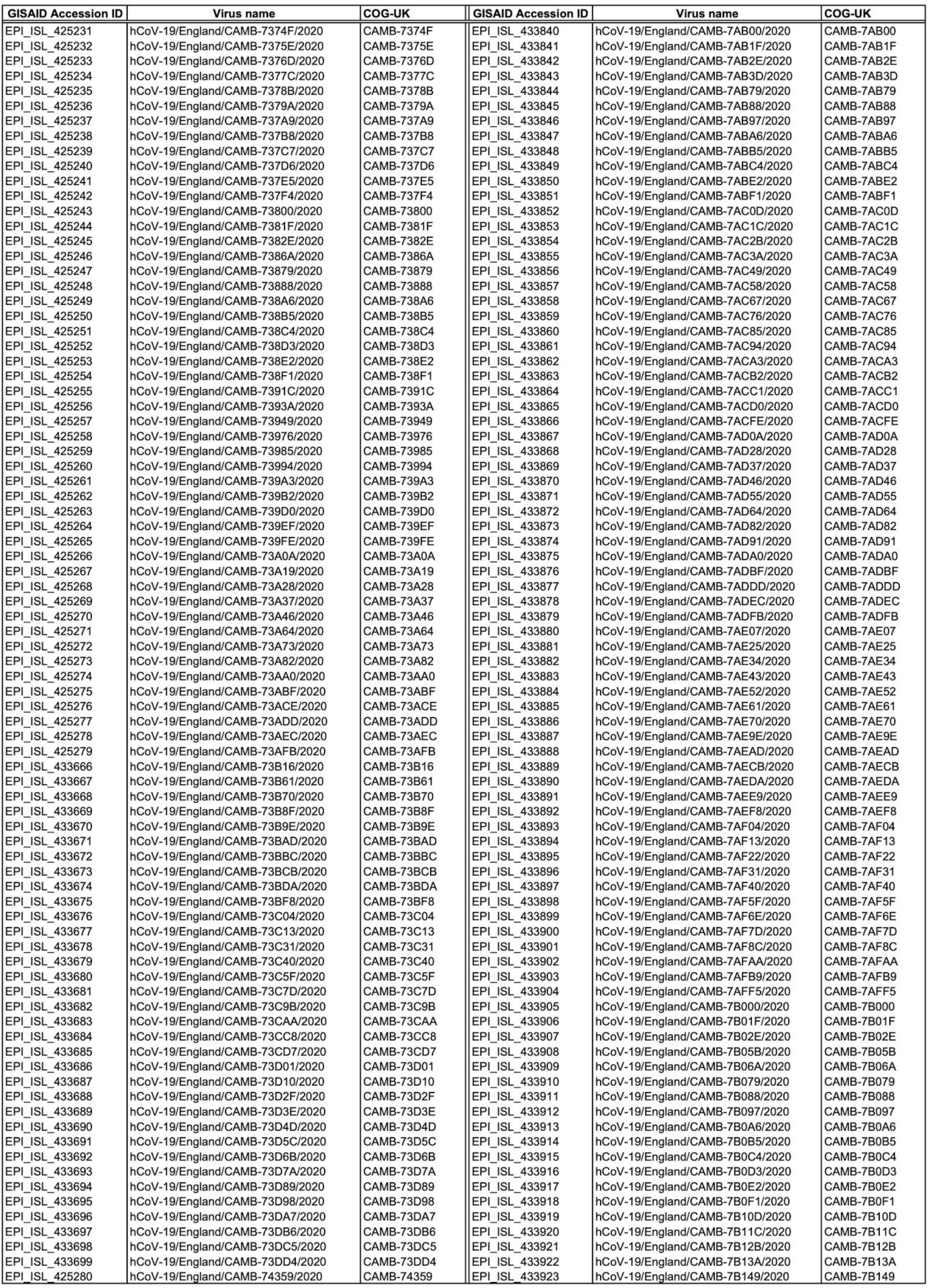
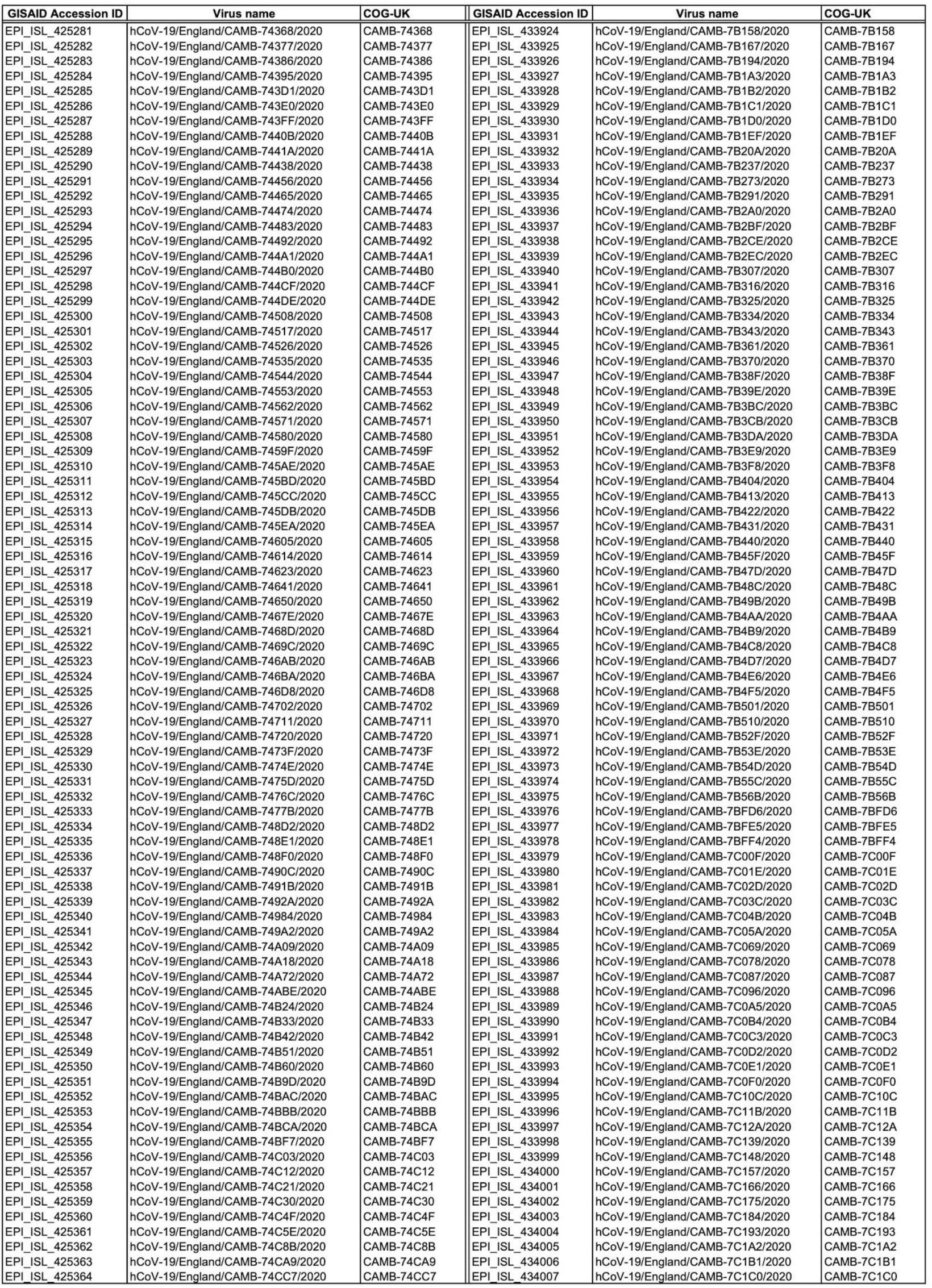
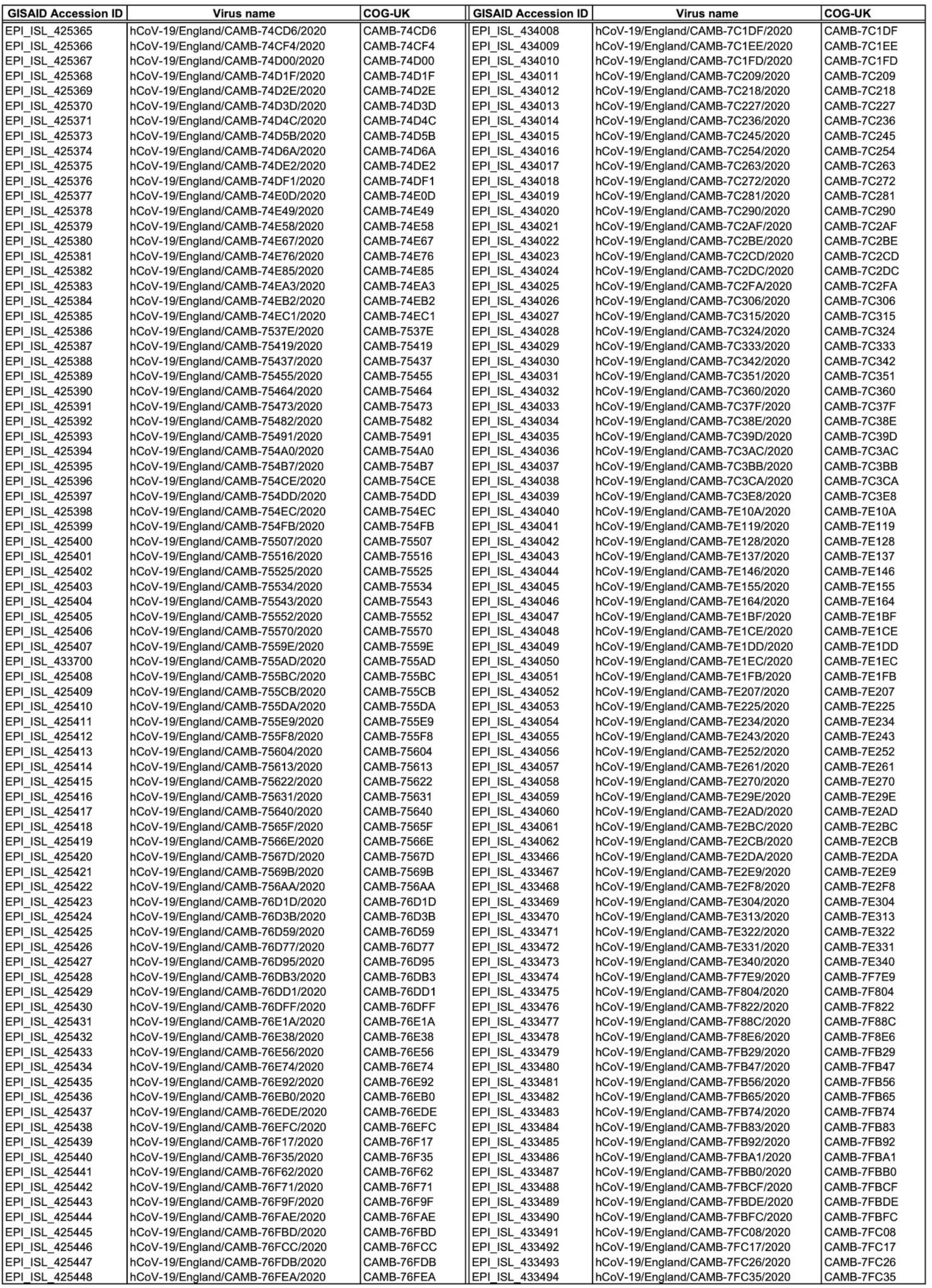
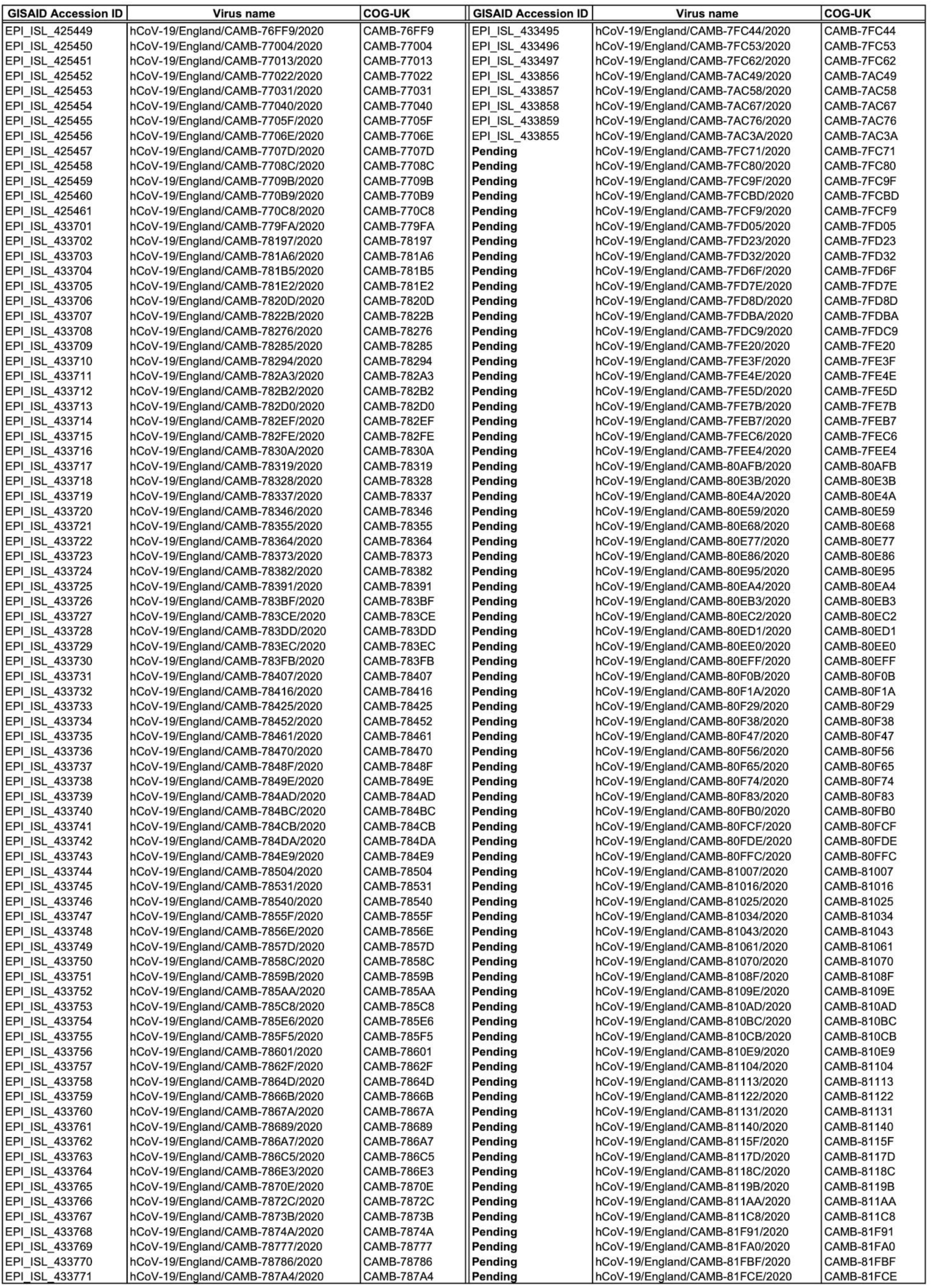
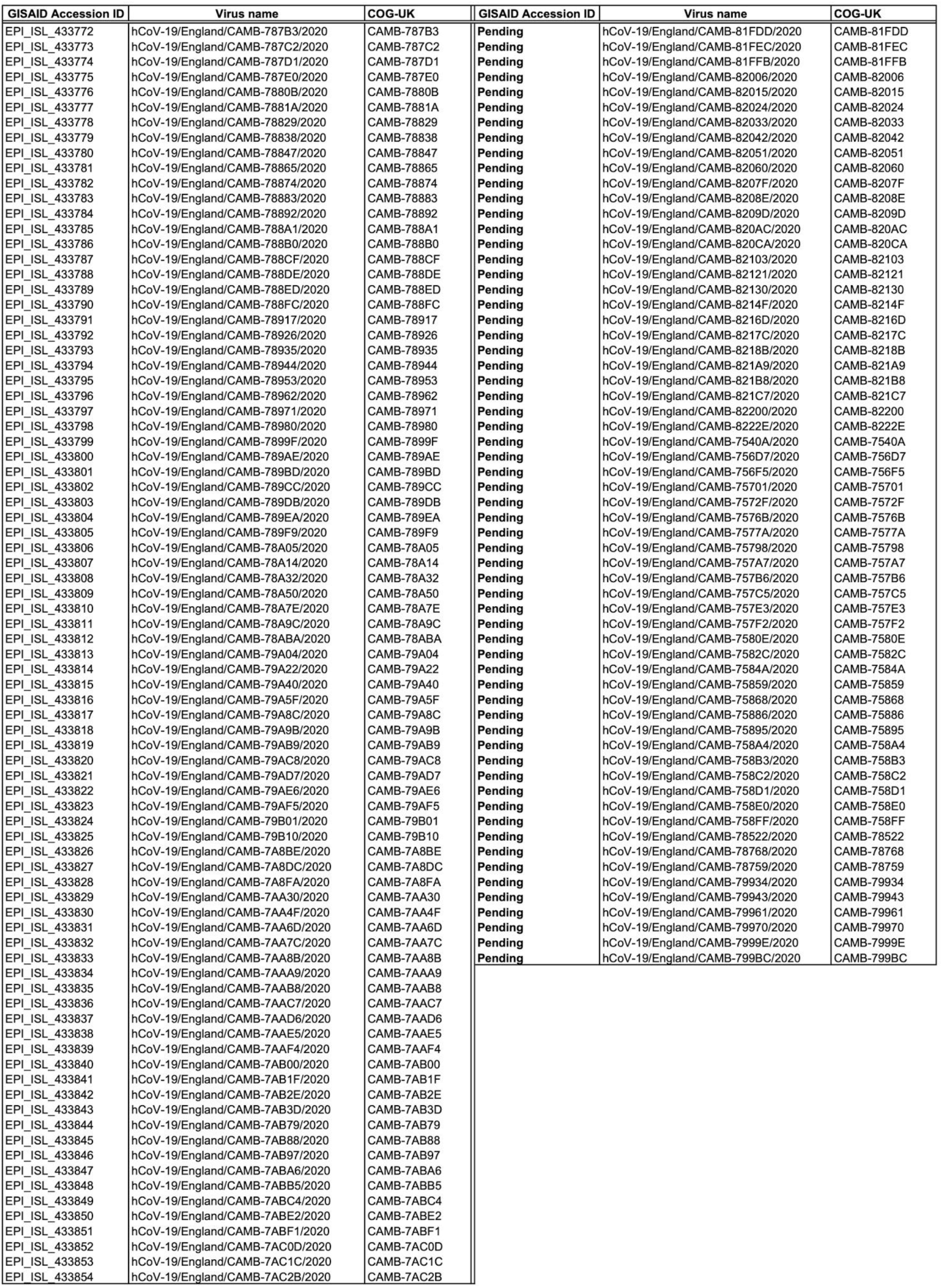
Table of sequences / accession numbers.

